# Optimizing Vaccine Allocation to Combat the COVID-19 Pandemic

**DOI:** 10.1101/2020.11.17.20233213

**Authors:** Dimitris Bertsimas, Joshua Ivanhoe, Alexandre Jacquillat, Michael Li, Alessandro Previero, Omar Skali Lami, Hamza Tazi Bouardi

## Abstract

The outbreak of COVID-19 has spurred extensive research worldwide to develop a vaccine. However, when a vaccine becomes available, limited production and distribution capabilities will likely lead to another challenge: who to prioritize for vaccination to mitigate the near-end impact of the pandemic? To tackle that question, this paper first expands a state-of-the-art epidemiological model, called DELPHI, to capture the effects of vaccinations and the variability in mortality rates across subpopulations. It then integrates this predictive model into a prescriptive model to optimize vaccine allocation, formulated as a bilinear, non-convex optimization model. To solve it, this paper proposes a coordinate descent algorithm that iterates between optimizing vaccine allocations and simulating the dynamics of the pandemic. We implement the model and algorithm using real-world data in the United States. All else equal, the optimized vaccine allocation prioritizes states with a large number of projected cases and sub-populations facing higher risks (e.g., older ones). Ultimately, the optimized vaccine allocation can reduce the death toll of the pandemic by an estimated 10–25%, or 10,000–20,000 deaths over a three-month period in the United States alone.

**Highlights:**

– This paper formulates an optimization model for vaccine allocation in response to the COVID-19 pandemic. This model, referred to as DELPHI–V–OPT, integrates a predictive epidemiological model into a prescriptive model to support the allocation of vaccines across geographic regions (e.g., US states) and across risk classes (e.g., age groups).
– This paper develops a scalable coordinate descent algorithm to solve the DELPHI–V–OPT model. The proposed algorithm converges effectively and in short computational times. Therefore, the proposed approach can be implemented efficiently, and allows extensive sensitivity analyses for scenario planning and policy analysis.
– Computational results demonstrate that optimized vaccine allocation strategies can curb the death toll of the COVID-19 pandemic by an estimated at 10–25%, or 10,000–20,000 deaths over a three-month period in the United States alone. These results highlight the critical role of vaccine allocation to combat the COVID-19 pandemic, in addition to vaccine design and vaccine production.

## 1 Introduction

In just a few months, the entire world has been upended by the COVID-19 pandemic, a severe acute respiratory disease caused by the SARS-CoV-2 coronavirus. First identified in Wuhan, China (Hui et al. 2020), the disease has spread rapidly, growing into a public health emergency on January 30, 2020 and a pandemic on March 11, 2020, according to the World Health Organization. As of August 2020, over 25 million cases and 800,000 deaths have been reported globally (Johns Hopkins University 2020). To curtail the viral spread of SARS-CoV-2, most governments have implemented non-pharmaceutical interventions spanning social distancing, self-isolation and travel restrictions, culminating in partial to full lockdowns in many parts of the world. As a result, the pandemic has also engendered unprecedented socio-economic strain, leading to growing unemployment and a looming recession in some of the world’s largest economies.

It is therefore of upmost priority for governments, healthcare providers and the general public to combat the COVID-19 pandemic. New non-pharmaceutical interventions continue to be designed and implemented to “flatten the curve” of the disease, prevent overflows in the healthcare system and allow more time to find new clinical solutions (Singh et al. 2020). At the same time, new clinical solutions are actively being researched and developed. These broadly fall into three categories: therapeutics, diagnostics, and vaccination (see Zhang et al. 2020, for a review). Therapeutics can alleviate the death toll and other health impacts among infected patients. Diagnostics can detect the presence of the virus or antibodies to prevent the near-term spread of the disease. Vaccines, on the other hand, intervene at the upstream level to immunize patients against the virus. Unsurprisingly, many pharmaceutical companies and academic institutions have been heavily investing into vaccine research: as of August 2020, over 100 experimental SARS-CoV-2 vaccines have been developed globally, 11 of which are in latest phases of clinical trials—an unprecedented speed for vaccine development (Lurie et al. 2020, Graham 2020).

Unfortunately, discovering and developing a vaccine for COVID-19 is just the beginning—it may take months to produce, distribute, and administer it at scale. Indeed, vaccine manufacturing involves tremendous complexity and high fixed costs (Plotkin et al. 2017), which will lead to a gradual ramp up in vaccine production capacity. And then, vaccine distribution will require the fast development of complex supply chains and numerous administration sites across the globe. Consequently, vaccines will not be available immediately to everyone, not withstanding the likely reality that a significant fraction of the population may not wish to get vaccinated (Cornwall 2020). So, how can policy makers determine who should be prioritized for vaccination to mitigate the near-end impact of the pandemic?

This paper addresses this question by developing a novel analytics-based approach to vaccine allocation. Obviously, vaccine allocation is highly complex, both quantitatively and qualitatively. For instance, there is a strong consensus that healthcare workers and other essential workers need to be prioritized. Moreover, vaccine allocation raises a number of ethical issues (Emanuel et al. 2020). Keeping these considerations in mind, this paper strives to provide data-driven evidence to guide vaccine allocation at a higher level, with the ultimate goal of utilizing available vaccines as effectively as possible to alleviate the death toll of the pandemic.

To this end, we leverage a recent compartmental epidemiological model called DELPHI (Differential Equations Lead to Predictions of Hospitalizations and Infections), which extends Susceptible-Exposed-Infected-Recovered (SEIR) models to capture critical drivers of the COVID-19 pandemic: (i) under-detection due to limited testing, (ii) governmental and societal response to the pandemic, and (iii) declining mortality rates. The DELPHI model has been fitted from historical data at the country level, at the state level in the United States, and at the province level in a few other countries. The DELPHI forecasts have been matching the number of detected cases and deaths with high accuracy (Li et al. 2020), and have thus been incorporated into the forecasts from the US Center for Disease Control (2020a).

In this paper, we integrate the (predictive) DELPHI model into a (prescriptive) optimization model for vaccine allocation. We first propose an extension of DELPHI, referred to as DELPHI–V, to capture the effects of vaccinations on the dynamics of the pandemic. The DELPHI–V model also disaggregates the dynamics at the subpopulation level to reflect disparities in mortality rates across age groups (Guan et al. 2020, Goyal et al. 2020, Petrilli et al. 2020), which are critical drivers of effective vaccination strategies. We then formulate an optimization model, referred to as DELPHI–V–OPT, which optimizes the number of vaccines allocated to each region and age group at each time period, over an extended planning horizon, to minimize the death toll of the pandemic.

From a technical standpoint, the DELPHI–V–OPT model relies on time discretization to embed the system of ordinary differential equations governing the DELPHI– V dynamics into an optimization model. The model is formulated as a bilinear (non-convex) optimization model. In this problem, the non-linearities and non-convexities stem from the SEIR dynamics at the core of the predictive DELPHI–V model, in which the number of new cases is a bilinear function of the susceptible and infected populations. We propose a coordinate descent algorithm to solve the DELPHI–V–OPT model efficiently in realistic large-scale settings. Starting from a baseline vaccine allocation, the algorithm iterates, until convergence, between optimizing the vaccine allocation solution (for given dynamics of the pandemic) and simulating the dynamics of the pandemic (for a given vaccine allocation strategy).

We implement the proposed modeling and algorithmic approach using real-world data in the United States from the New York Times (2020), the US Census Bureau (2020b), and the US Center for Disease Control (2020b). We leverage parameter calibration from the DELPHI model in each US state. One challenge, however, lies in developing realistic and consistent estimates for mortality rates in each state and in each age group— the DELPHI model estimates mortality rates in each state every day, while the US Center for Disease Control (2020b) reports mortality rates in each age bracket. To solve this discrepancy, we formulate an optimization model that interpolates these two pieces of information, while ensuring consistency with broader demographic information. The model is formulated as another bilinear, non-convex optimization problem.

Experimental results yield three main takeaways. First, the coordinate descent algorithm terminates in a few iterations and yields stable vaccine allocation solutions, regardless of the initialization. These strong convergence properties enable the efficient implementation of the proposed optimization model, as well as extensive sensitivity analyses for scenario planning and policy analysis. Second, our optimization approach results in significantly lower numbers of deaths than a proportional allocation benchmark (which allocates vaccines proportionally to the size of each subpopulation). Specifically, we estimate that the optimized vaccine allocation solution can reduce the death toll of the pandemic by 10–25%, or 10,000–20,000 deaths over a three-month period in the United States alone. Moreover, these benefits are highly robust to misspecifications and fluctuations in the DELPHI parameters. Third, we uncover that optimized vaccine allocation is governed by two key drivers: (i) the near-term dynamics of the pandemic in each state; and (ii) the risk level of each age group. By resolving the resulting trade-offs between allocating vaccines to medium-risk populations in a “hot spot” vs. high-risk populations in a “better” state, the proposed optimization model provides effective decision-making tools to support vaccine allocation planning.

In summary, this paper makes three contributions. From a modeling standpoint, it formulates an original optimization model for vaccine allocation, DELPHI–V– OPT, that integrates a state-of-the-art epidemiological model to mitigate the impact of the pandemic at a disaggregate subpopulation level. From a computational standpoint, it develops a scalable coordinate descent algorithm, which converges effectively and in short runtimes. From a practical standpoint, it demonstrates that optimized vaccine allocation strategies can curb the death toll of COVID-19 by a sizeable amount—highlighting the critical role of vaccine allocation besides vaccine design and vaccine production. We are in discussion with the manufacturing and supply chain divisions of a major pharmaceutical company to inform vaccine distribution planning based on the outputs of the model.

## 2 Literature review

Many pharmaceutical companies and academic institutions have explored different technologies toward a SARS-CoV-2 vaccine, spanning (i) inactivated or live-attenuated virus vaccines, which induce an immune response from weakened or killed pathogens (used by the University of Hong Kong and Codagenix, for instance); (ii) viral vector vaccines, which exploit non-replicating adenoviruses to deliver an antigenic element (used by the Chen Wei group and Johnson and Johnson, for instance); (iii) subunit vaccines, which use a minimal structural component of a pathogen such as a protein (used by Novavax and Clover Biopharmaceuticals, for instance); (iv) nucleic acid vaccines, which deliver DNA or mRNA of viral proteins, such as the spike protein (used by Pfizer and Moderna, for instance). For a review of latest progress toward a COVID-19 vaccine, we refer to Shin et al. (2020) and Florindo et al. (2020).

From an operational standpoint, a vast literature studies the optimization of the management of vaccine supply chains (see Duijzer et al. 2018, Lemmens et al. 2016). A first area involves optimizing vaccine composition for maximal effectiveness (Wu et al. 2005, Kornish and Keeney 2008, Cho 2010, Robbins and Jacobson 2011). A second area involves optimizing vaccine production to manage supply-side and demand-side uncertainties (Federgruen and Yang 2009, De Treville et al. 2014) and to design market coordination mechanisms that address incentive misalignments between manufacturers and end users (Chick et al. 2008, Arifoğlu et al. 2012). Next, vaccine allocation optimizes the management of a vaccine stockpile—this is the focus of this paper. Last, vaccine distribution focuses the final phase of the planning process by optimizing distribution networks (Kaufmann et al. 2011), inventory management (Jacobson et al. 2006a), and dispensing operations (Aaby et al. 2006). Most of this research focuses on planning for predictable and repeatable epidemics, such as seasonal influenza. For less predictable epidemics, such as pandemic influenza, studies have suggested advanced planning interventions, including stockpiling (Jacobson et al. 2006b) and anticipatory vaccination (Arinaminpathy et al. 2012). Unfortunately, these approaches are not readily applicable to the case of a new disease, such as COVID-19. The literature remains scarce when it comes to vaccine management in response to sudden outbreaks (Duijzer et al. 2018).

Focusing on vaccine allocation, a first line of research studies inter-country competition. Sun et al. (2009) develop a game-theoretic framework for vaccine stockpile management, in which each country faces a trade off between keeping vaccines to protect its own population and sharing them with highly infected countries to prevent the spread of the outbreak. In a dynamic setting, Mamani et al. (2013) find that a lack of coordination leads to a shortage of vaccines in some regions and to an excess in others, and propose a contract to alleviate these inefficiencies.

In contrast, our paper considers a problem of centralized allocation of vaccines within a population. Using infection models based on heterogeneous mixing of populations, early studies established the importance of partitioning the population into risk classes (e.g., age groups) to accurately estimate the impact of an epidemic (Watson 1972, Elveback et al. 1976, Longini Jr et al. 1978). Emanuel and Wertheimer (2006) propose a life-cycle model that prioritizes the most valuable sub-populations. Some studies have leveraged the epidemiological dynamics of the disease to optimize the allocation of vaccines among age groups, with a focus on influenza vaccination. Results suggest prioritizing at-risk populations (Patel et al. 2005, Chowell et al. 2009) or active agents who can spread the disease fastest, such as school children (Dushoff et al. 2007, Basta et al. 2009, Medlock and Galvani 2009, Matrajt and Longini Jr 2010, Lee et al. 2012, Matrajt et al. 2013). In a multiregion setting, results suggest that vaccines should be allocated to the most infected regions and to the regions affected the latest by the epidemic (Araz et al. 2012, Keeling and Shattock 2012).

All these studies integrate SEIR or similar epidemiological models into simple optimization routines based on scenario analysis, enumeration, simulation, or simple heuristics (e.g., genetic algorithms). Similar simulation-based heuristics have been proposed in the operations research community (Uribe-Sánchez et al. 2011, Teytelman and Larson 2013). In contrast, Tanner et al. (2008) propose a chance-constrained optimization approach that optimizes vaccine allocation while ensuring that the post-vaccination reproduction number is lower than one with high probability, under uncertainty in the dynamics of disease propagation. Using connections to the newsvendor problem, Yarmand et al. (2014) formulate an efficient two-stage stochastic programming model to optimize first-stage vaccine allocation planning and a second-stage recourse to distribute additional doses where the epidemic has not been contained. They model the dynamics of disease propagation by means of a stochastic SEIR model, and define scenarios using Monte Carlo simulation.

This paper expands this overall body of work in three major ways. First, we optimize vaccine allocation across regions and across risk classes (e.g., age groups) over an extended planning horizon, using a data-driven procedure to estimate mortality rates dynamically at the subpopulation level. Second, we leverage a recent SEIR-inspired epidemiological model that captures dynamics specific to the COVID-19 pandemic, such as under-detection, different waves of governmental response, and declining mortality rates over time. Third, we propose a formal optimization approach and a coordinate descent algorithm to embed the predictive epidemiological model into a prescriptive vaccine allocation model, as opposed to relying on enumeration, simulation or simplified heuristics.

## 3 Model formulation

Our model optimizes vaccine allocation to minimize the death toll of the COVID-19 pandemic. We capture the dynamics of the pandemic by means of an epidemiological model, called DELPHI, which forecasts the number of detected cases, hospitalizations and deaths for each country, each US state, and each province of other large countries (Li et al. 2020).

Before proceeding further, let us highlight two critical aspects of the COVID-19 pandemic:

– *Variable mortality rate across risk classes* (Guan et al. 2020, Goyal et al. 2020, Petrilli et al. 2020): One of the primary sources of variation in mortality rates is age. The US Center for Disease Control (2020b) reports that the mortality rate among Americans aged 70 and over is two orders of magnitude greater than for those aged 30 and under. We partition the population into *risk classes*, defined as homogeneous groups with comparable health characteristics. We consider age-based risk classes in our experiments, but other categorization could be used (e.g., based on comorbidities).
– *Variable severity by region*: The outbreak of the COVID-19 pandemic has impacted different geographic areas differently (Garg 2020, Vahidy et al. 2020). For instance, in March 2020, the New York state accounted for over 40% of nationwide COVID-19 deaths, despite representing around 6% of the US population (New York Times 2020). We define a *region* as a geographic entity with similar disease propagation dynamics. We consider in our experiments 51 regions, corresponding to the 50 US states plus the District of Columbia.

We refer to as a *subpopulation* the group of people belonging to a given risk class in a given region. Our prescriptive model designs a targeted vaccine allocation at the subpopulation level (for each risk class in each region). However, the predictive DELPHI model is fitted at the region level (across all risk classes) but not at the subpopulation level. In other words, the predictive DELPHI model forecasts the number of aggregate cases, hospitalizations and deaths in each region—not the number of cases, hospitalizations and deaths in each age group within each region. This is justified by them lack of disaggregated information in publicly available case counts, but also by the limited number of observations (fitting the DELPHI model at the subpopulation level would significantly increase the variance of the estimates). Accordingly, we model the dynamics of infections at the region level but disaggregate the dynamics of mortality at the subpopulation level.

We first briefly review the original DELPHI model— for further details, we refer to Appendix A and to Li et al. (2020). We then extend the DELPHI model to capture the effects of vaccinations and variability in mortality rates across risk classes. We refer to the resulting DELPHI model with vaccinations as DELPHI– V. Last, we embed the DELPHI–V model into an original linear programming formulation to optimize vaccine allocation, referred to as DELPHI–V–OPT.

### 3.1 The DELPHI model: Forecasting the dynamics of the COVID-19 pandemic

DELPHI is a compartmental epidemiological model, which extends the widely used SEIR model to account for effects germane to the COVID-19 pandemic. The model is governed deterministically by a system of ordinary differential equations (ODEs) involving 11 states: susceptible (*S*), exposed (*E*), infectious (*I*), undetected cases who will recover (*U*^*R*^) or die (*U*^*D*^), detected hospitalized cases who will recover (*H*^*R*^) or die (*H*^*D*^), detected quarantined cases who will recover (*Q*^*R*^) or die (*Q*^*D*^), recovered (*R*) and deceased (*D*). The separation of undetected, quarantined and hospitalized cases into recovering and dying states enables separate fitting of recoveries and deaths from the data.

DELPHI differs from other COVID-19 forecasting models (see, e.g. Kissler et al. 2020) by capturing three key elements of the pandemic:

– **Under-detection**: Many cases remain undetected due to limited testing, asymptomatic carriers, and detection errors. Ignoring them would underestimate the scale of the pandemic. The DELPHI model captures them through the *U*^*R*^ and *U*^*D*^ states.
– **Governmental and societal response**: Social distancing policies limit the spread of the virus. Ignoring them would overestimate the scale of the pandemic. However, if restrictions are lifted prematurely, a resurgence may occur. We define a governmental and societal response function *γ*(*t*), which modulates the infection rate and is parameterized as follows:

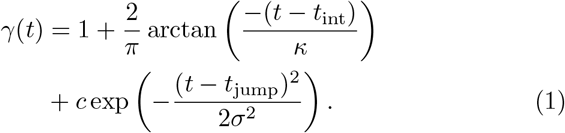 This parameterization encompasses four phases (Figure 1). In Phase I, most activities continue normally as people adjust their behaviors. This is followed by a sharp decline in the infection rate during Phase II as the policies get implemented. The parameters *t*_int_ and *κ* can be interpreted as the start time and the strength of this response. In Phase III, the decline in the infection rate reaches saturation. The epidemic then experiences a resurgence of magnitude *c* in Phase IV, due to relaxations in governmental restrictions and in social behaviors. This is counteracted at time *t*_jump_, when restrictions are re-implemented, with *σ* controlling the duration of this second wave.
– **Declining mortality rates**: The mortality rate of COVID-19 has been declining through the pandemic, due to a better detection of mild cases, enhanced care for COVID-19 patients, and other factors. We model the mortality rate as a monotonically decreasing function of time:

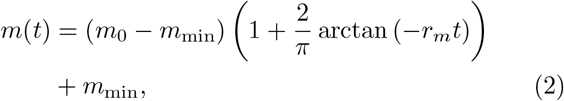

where *m*_0_ is the initial mortality rate, *m*_min_ is the minimum mortality rate and *r*_*m*_ is a decay rate.

**Fig. 1:**
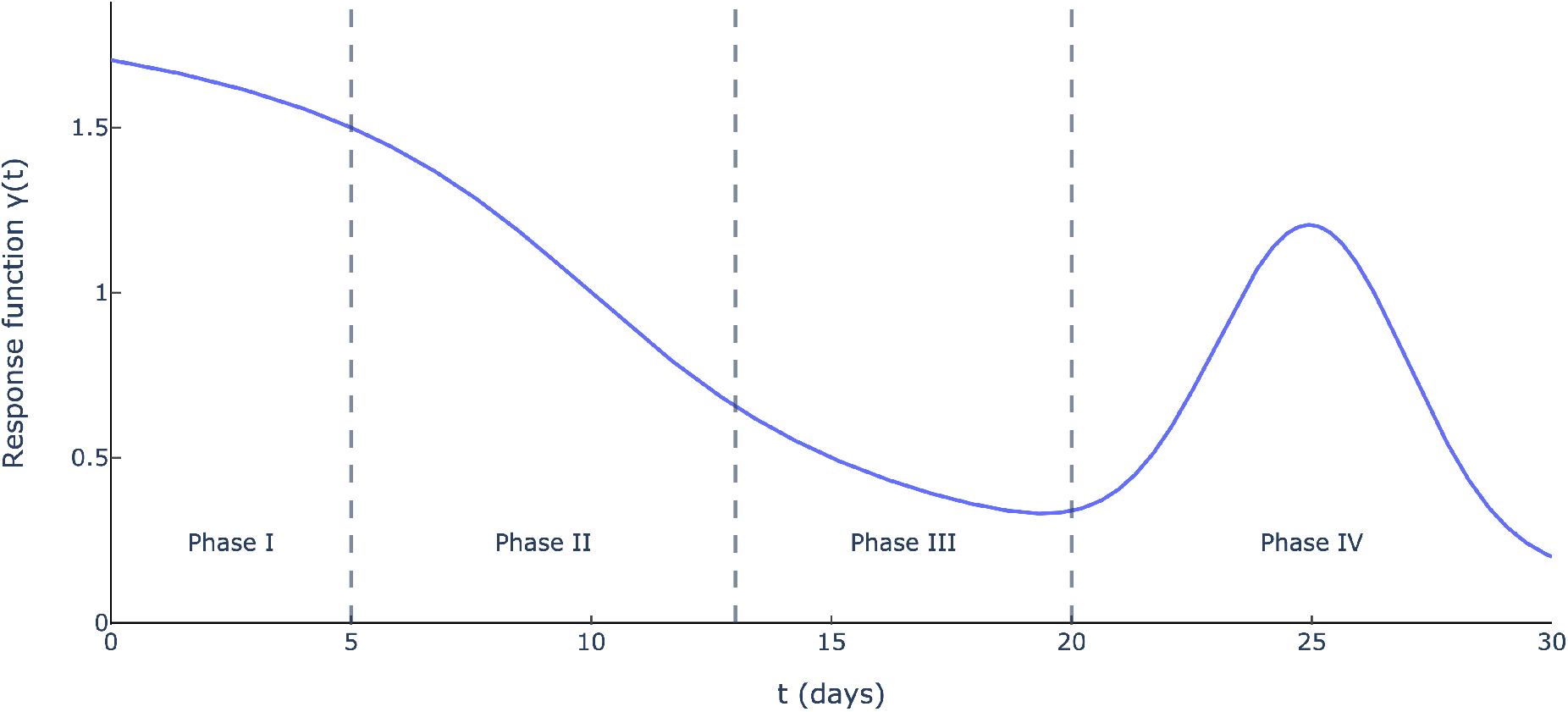
Governmental and societal response function *γ*(*t*) (*κ* = 5, *t*_int_ = 10, *c* = 1, *t*_jump_ = 25 and *σ* = 2).

Ultimately, DELPHI involves 16 parameters that define the transition rates between the 11 states. We calibrate 7 of them from a database on clinical outcomes (Bertsimas et al. 2020). Using non-linear optimization, we estimate the other 9 parameters from historical data on the number of cases and deaths in each region. We provide details on the fitting procedure in Appendix A.

Since its inception in March 2020, DELPHI has been extensively tested and validated against real-world data, using a backtesting procedure. Table 1 reports the median Mean Absolute Percentage Error (MAPE) across states in each US region, defined by the US Census Bureau (2020a) and comprising 9–17 states each. Specifically, we fit the model’s parameters using data up to July 15 (the starting point of our vaccine allocation experiments), we build projections for a duration of 15 days, 30 days and 45 days, and we evaluate the MAPE against actual observations.

**Table 1:** Median out-of-sample MAPE aggregated per US region (prediction made on July 15th, 2020 MAPE evaluated over 15 days, 30 days, and 45 days).

| Region | # States | Cases |  |  | Deaths |  |  |
| --- | --- | --- | --- | --- | --- | --- | --- |
|  |  | 15 d. | 30 d. | 45 d. | 15 d. | 30 d. | 45 d. |
| Midwest | 12 | 11.4% | 16.4% | 24.2% | 6.3% | 7.2% | 9.5% |
| Northeast | 9 | 2.6% | 4.7% | 7.3% | 1.9% | 2.6% | 3.0% |
| South | 17 | 8.4% | 8.7% | 13.9% | 13.2% | 13.1% | 12.0% |
| West | 13 | 9.1% | 14.4% | 17.2% | 12.2% | 12.6% | 16.7% |
| All | 51 | 8.4% | 12.0% | 16.6% | 8.7% | 8.9% | 9.4% |

Note that the MAPE remains relatively small. For instance, two-week predictions result in a nation-wide median MAPE of 8–9% for both cases and deaths. Over the period considered, the model performed well in the Northeast but worse in the Midwest, where the dynamics of the pandemic were more volatile. Given the high level of uncertainty and variability in the disease’s spread, these results suggest adequate out-of-sample performance, which enables us to leverage the DELPHI model in our vaccine allocation optimization. Moreover, the MAPE increases as the prediction window gets longer. In other words, the DELPHI model captures well the short-term dynamics of the pandemic but is less accurate on the medium-to long-term dynamics. In practice, we can thus run our vaccine allocation optimization model on a rolling basis, where we project the DELPHI predictions over an extended planning horizon but only implement the short-term outputs of the model—iterating over time to leverage the latest data in the DELPHI predictions.

### 3.2 The predictive DELPHI–V model

Capturing the effects of vaccination. We now augment the DELPHI model to capture the impact of vaccinations. We refer to the new model as DELPHI–V. Specifically, we implement two main changes to the original DELPHI model:

1. Impact of vaccinations on the dynamics of the pandemic. The primary, direct effect of vaccinations is that a fraction of vaccinated people will be immune to the disease (based on the vaccine’s effectiveness). A second, indirect effect is that reducing the number of infections will lead to a slower spread of the disease among non-vaccinated people. To capture these effects, we create a new state in the model, referred to as “immune” and denoted by *M*.
2. Disparate impacts of the disease and vaccinations across risk classes. We replicate the 12 states (the 11 original states plus the new immune state) for each risk class.

Figure 2 shows the flow diagram of the DELPHI–V model, with two risk classes (indexed by *k* = 1, 2 and indicated via subscripts). For exposition purposes, we omit dependencies on the region (since the DELPHI–V model is fitted in each region independently).

**Fig. 2:**
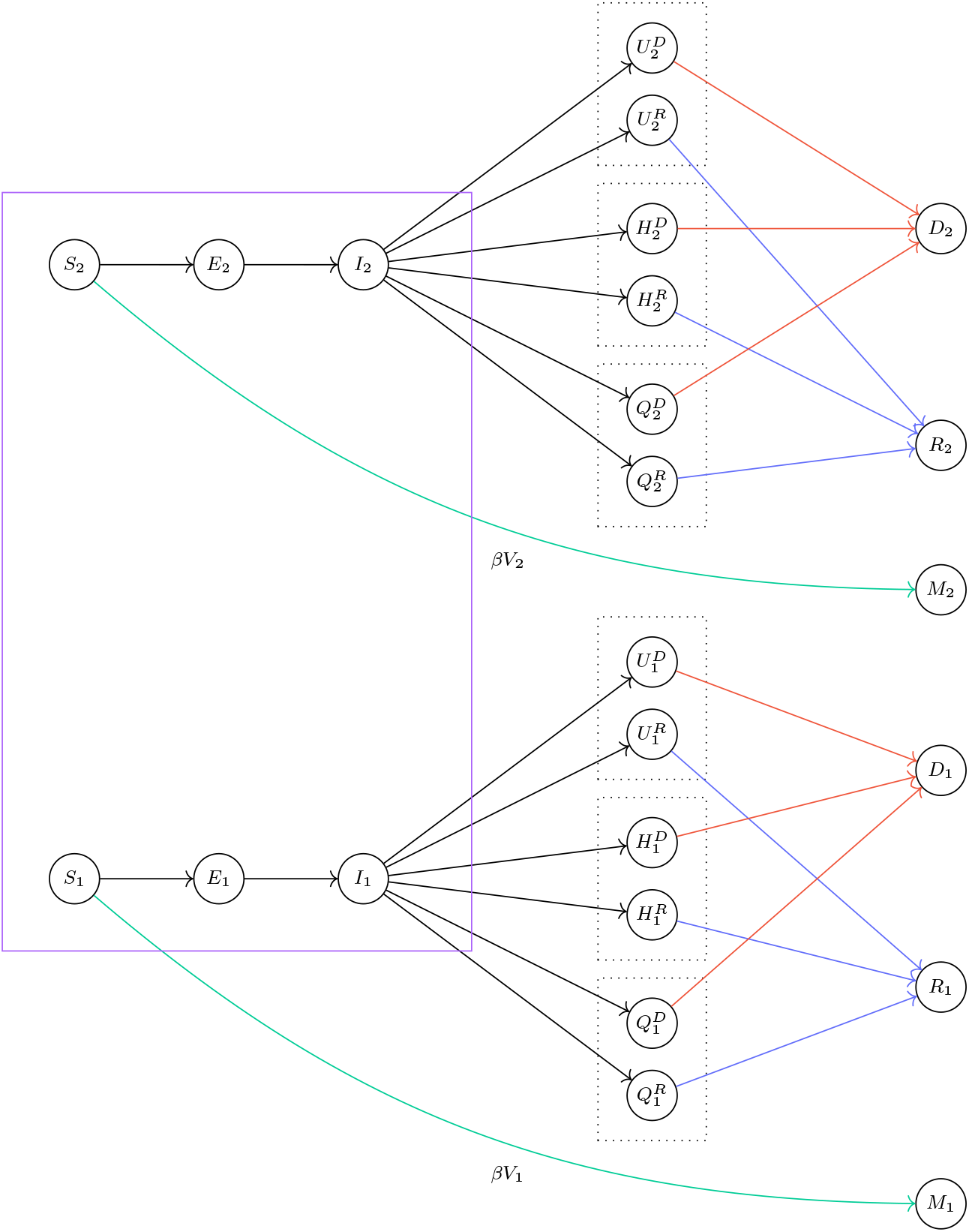
Simplified flow diagram of the DELPHI–V model.

Let *V*_*k*_(*t*) denote the population mass from risk class *k* that gets vaccinated at time *t*, and let *β* ∈ (0, 1] denote the vaccine’s effectiveness. For simplicity, we assume that the effects of vaccines (hence, the transitions from the susceptible state to the immune state) are instantaneous. Moreover, we assume that the vaccine has no effect when it fails to immunize the patient (i.e., no partial benefit and no side effect). Then, at each time *t*, a mass *βV*_*k*_(*t*) of people transitions from the susceptible state *S*_*k*_ to the immune state *M*_*k*_, and the remaining mass (1 − *β*)*V*_*k*_(*t*) remains in the susceptible state. All other transitions shown in Figure 2 are consistent with the original DELPHI model.

The DELPHI–V model is then governed by the following ODE system:

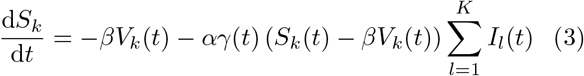

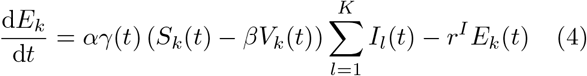

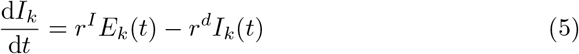

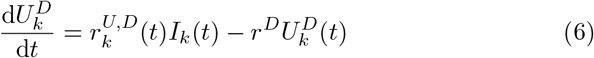

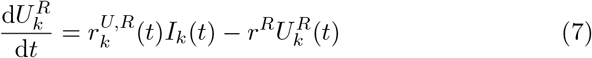

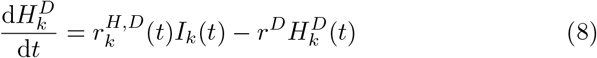

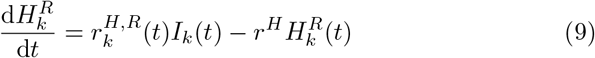

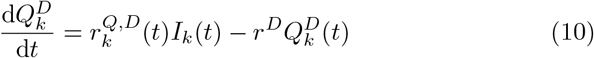

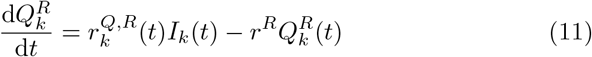

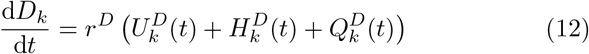

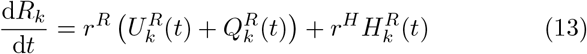

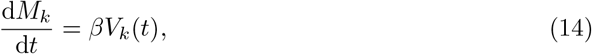

– *α* is the nominal infection rate;
– *γ*(*t*) : ℝ_+_ → ℝ_+_ is the governmental and societal response function (Figure 1);
– *r*^*I*^, *r*^*d*^, *r*^*D*^, *r*^*R*^ and *r*^*H*^ are the progression rate, the detection rate, the death rate, the recovery rate for non-hospitalized patients, and the recovery rate for hospitalized patients;
– 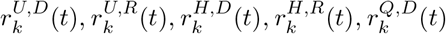 and 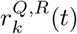 capture the disaggregated detection, hospitalization and death rates—accounting for the probability of detection, the probability of hospitalization, and the mortality rate given in Equation (2). By design, these six rates sum up to *r*^*d*^. The notations reflect their dependencies over time as well as their dependencies across risk classes, thus capturing the disparate effects of the pandemic over time and across subpopulations.

As noted earlier, the dynamics of exposure and infection depend on the total number of infected people in a state, as opposed to the number of infected people in any particular risk class. Accordingly, DELPHI–V captures these interdependencies across risk classes— indicated by the purple rectangle in Figure 2, and reflected in the terms 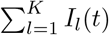 in Equations (3)–(4).

Given initial conditions for each of the 12 states, these equations uniquely determine the evolution of this system over time—for a given vaccine allocation. Next, we optimize vaccine allocation to minimize the overall impact of the pandemic—estimated by DELPHI–V.

## 3.3 The prescriptive DELPHI–V–OPT model: Optimizing vaccine allocation

The vaccine allocation model takes as inputs (i) epidemiological data for each region; (ii) clinical information on the vaccine; and (iii) data on the availability of the vaccine. On the epidemiological side, we import all parameters calibrated from the DELPHI–V model. On the clinical side, we encapsulate vaccine characteristics through the effectiveness parameter *β*. On the vaccine availability side, we consider a daily budget of vaccines that can be allocated across the entire population. Note that this budget can vary over time to reflect the capacity of the underlying supply chains.

We proceed by time discretization to formulate the optimization model and retain tractability. This reduces to solving the system of ODE equations given in Equations (3)–(13) by a forward difference scheme. We denote by *Δt* the discretization unit (e.g., 1, 2 days).

Formally, we define a set of time periods, indexed by *t* ∈ 𝒯 = {1, …, *T}*; a set of regions, indexed by *j* ∈ ℳ = *{*1, *…, M*}; and a set of risk classes, indexed by *k* ∈ 𝒦 = *{*1, *…, K}*. The progression rate *r*^*I*^, the detection rate *r*^*d*^, the death rate *r*^*D*^ and the recovery rate *r*^*R*^ are constant across regions and risk classes, since they characterize clinical features of the disease. The same holds for *β*, which characterizes features of the vaccine. The nominal infection rate and the governmental and societal response depend on the region, reflecting geographical disparities in the spread of the pandemic. Accordingly, we now denote them by *α*_*j*_ and *γ*_*jt*_. We do not assume any differences in infection and transmission rates between the different risk classes. In contrast, we capture variations in mortality rates across risk classes (at the subpopulation level). We thus denote the disaggregated rates by 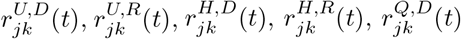 and 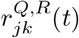. We also denote by *N*_*jk*_ the total population in region *j* ∈ ℳ and in risk class *k* ∈ 𝒦.

From these inputs, the model optimizes the number of vaccines allocated in each region, risk class and time period, which we capture by decision variables *V*_*jkt*_. In order to track the impact of vaccine allocation on the resulting dynamics of the pandemic, we create indirect variables, corresponding to eight states in the DELPHI– V model (we omit the states from patients who recover since they do not impact the objective function).

The vaccine allocation problem, referred to as (𝒫), is then formulated in Equations (16)–(31).

Equation (15) minimizes the number of deaths over the planning horizon, across all regions and risk classes. This includes all people in the absorbing state *D*, as well as the transient states *U*^*D*^, *H*^*D*^, and *Q*^*D*^, by period *T*. Equations (16)–(23) capture the dynamics of the DELPHI–V model in a discretized time space (Equations (3)–(13)). The remaining constraints capture practical considerations surrounding vaccine allocation:

– **Inter-regional capacity**: Due to restrictions in vaccine manufacturing and distribution networks, a limited number of vaccines can be allocated in each time period. Equation (24) ensures that the total number of vaccines allocated lies within the available budget in each time period.
– **Eligibility**: We prevent people from being vaccinated twice: a patient who has been vaccinated but remains susceptible cannot be vaccinated again. At period *t*, the number of eligible people in region *j* and in risk class *k* is denoted by 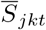 (an auxiliary decision variable). The number of people who have been vaccinated by period *t* in region *j* and in risk class *k* is equal to 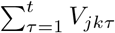, which is equal to 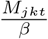 by Equation (23). Equation (25) then defines 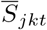 as the difference between the number of remaining susceptible people and the number of people vaccinated up to period *t*. Equation (26) ensures that the number of allocated vaccines does not exceed the eligible mass.
– **Safety**: It may be necessary to avoid vaccinating individuals with developing immune systems (e.g., young children) or weak immune systems (e.g., elderly people). Equation (27) states that no individual in an excluded risk class *k* ∈ *ε* may receive a vaccine, due to safety considerations.
– **Intra-regional capacity**: In addition to the overall budget per time period, each region is also constrained by its own ability to administer vaccines to its constituents. For example, even if 1 million vaccines were available in the United States per day, it would be unreasonable to expect the entire population of Wyoming to be vaccinated in a single day. Equation (28) bounds the total vaccines allocated to any region at a given time period by a fraction *f*_max_ of its population.
– **Inter-regional fairness**: From a pragmatic and ethical perspective, we promote a relatively equitable distribution of vaccines across regions. To be politically and socially viable, vaccine allocation must avoid neglecting any region, even if it is not a virus “hot spot”. This also promotes robustness in the solution, given that inter-regional transmission can occur in practice. Equation (29) ensures the allocation of vaccines to at least a fraction *f*_min_ of the eligible susceptible population entitled to a vaccine. For example, with a budget for vaccinating 1% of the population in a given time period and *f*_min_ = 0.5, each region is guaranteed to receive vaccines for 0.5% of its eligible susceptible population in that time period. Crucially, this still leaves at least (1 − *f*_min_)*b*_*t*_ vaccines available to allocate to the subpopulations with the most need.
– **Smoothness**: Large fluctuations in the number of vaccines allocated to each region from day to day would likely cause problems from a supply chain management perspective—both to deliver and to administer the vaccines. Equation (30) limits the change in any regional vaccine allocation to a fraction *δ* of its maximum capacity 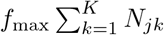.

### Model Structure

Problem (𝒫) is a non-linear programming model. The key complexity lies in the bilinear terms in Equations (16)–(17), which reflect the fact that the number of new infections result from the interactions between susceptible and infected populations—a key characteristic of all compartmental models based on SEIR dynamics. These bilinear terms result in non-linear, non-convex constraints, and thus in a highly challenging optimization model.

The latest Gurobi 9.0 release includes a solver for non-convex quadratic problems (Gurobi Optimization 2020). Yet, existing general-purpose technologies are limited to small-scale instances. In our setting, Problem (𝒫) includes 2*MKT* non-convex constraints each involving 2*K* bilinear terms, for a total of 4*MTK*^2^ bilinear terms. For context, a realistically-sized problem with *M* = 51 (50 US states plus Washington, D.C.), *K* = 6 (6 age groups) and *T* = 90 (a 3 month planning horizon with daily discretization) would result in 660,960 bilinear terms. Ultimately, Problem (𝒫) remains intractable with existing commercial solvers, motivating the development of a tailored algorithm to solve it in realistic settings.

## 4 Solution algorithm

We propose an iterative coordinate descent algorithm to solve Problem (𝒫) (Equations (15)–(31)), in order to

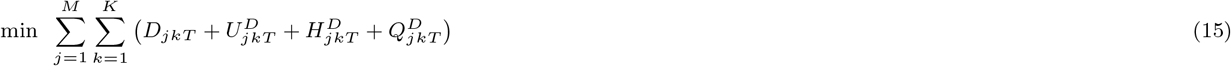

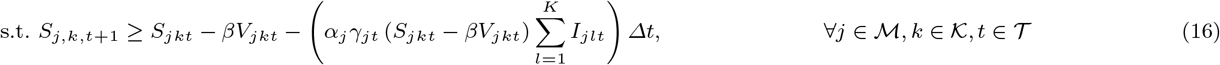

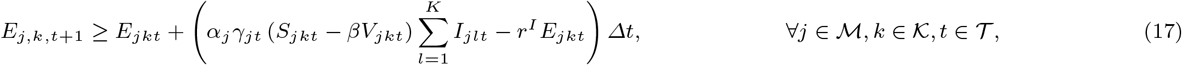

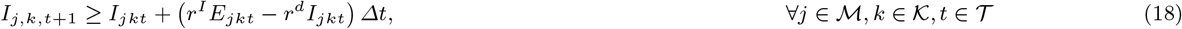

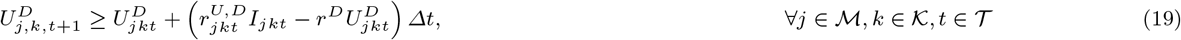

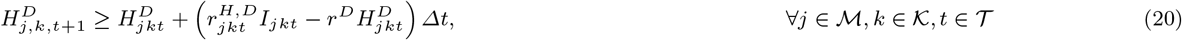

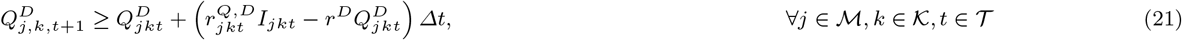

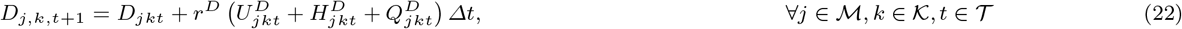

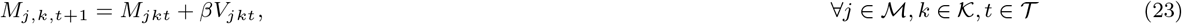

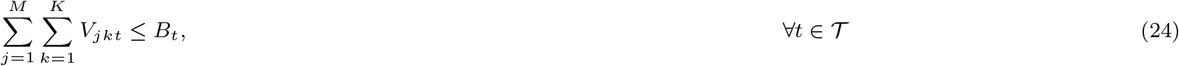

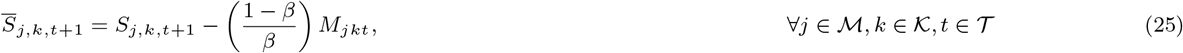

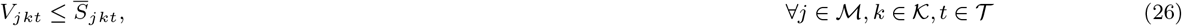

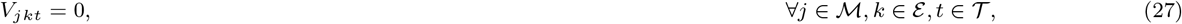

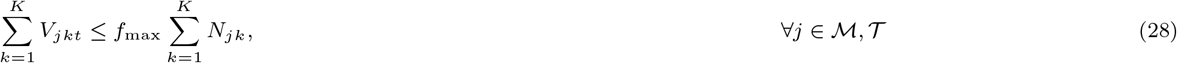

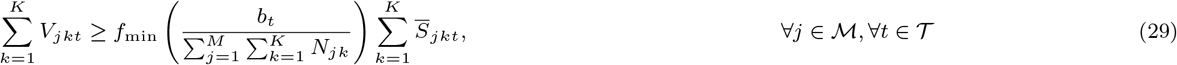

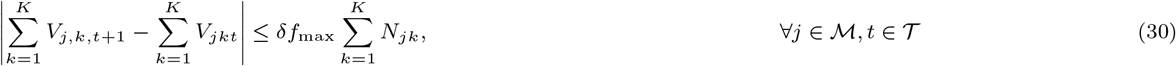

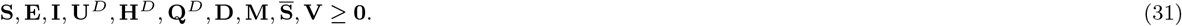

generate high-quality solutions in short computational times—consistent with practical requirements. We first describe the algorithm. We then present heuristics to generate initial feasible solutions, as well as a benchmark replicating reasonable policies that could be implemented in the absence of data-driven optimization.

### 4.1 Algorithm design

The key observation underlying our algorithm is that, aside from Equations (16)–(17), the objective function and all other constraints in (𝒫) are linear. Therefore, we proceed with a coordinate descent procedure that alternates between two modules: one that simulates the dynamics of the pandemic for a given vaccination allocation, and one that optimizes vaccination allocation given the infection dynamics. By fixing the bilinear terms one at a time, each of these two modules involves linear dynamics, which can be solved very efficiently. Specifically, the two modules are defined as follows:

1. **Simulate**: A vaccine allocation solution **V** uniquely determines a solution to DELPHI–V, which replicates the evolution of the pandemic from *t* = 0 to *t* = *T* (Section 3.2). This can be done by solving the ODE system (Equations (3)–(13)) using a forward difference scheme in a discretized time space, which terminates in 𝒪 (*MKT*) operations. Equivalently, we can fix the variable **V** and solve Equations (15)– (23). We denote the total infected population in region *j* and in time period *t* by 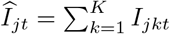. We refer to this procedure as `Simulate` (**V**).
2. **Optimize**: Given the infectious population estimates **Î**, we can approximate Equation (16) and (17) by the following linear constraints:

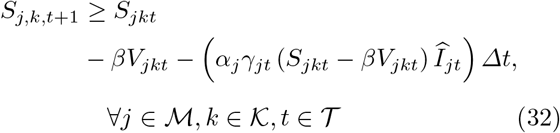

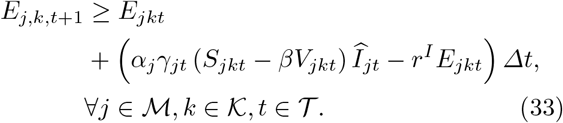

We also add the following regularization constraint, to bound the error of our approximation by a prespecified error tolerance *ε*:

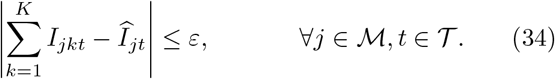

This constraint ensures that the vaccine allocation solution does not lead to infection dynamics that are vastly different that those estimated with the previous solution. The parameter *ε* can be viewed as an “exploration tolerance” that encapsulates the exploration/exploitation trade-off in the algorithm. If *ε* is too small, the algorithm may exhibit slow convergence or reach a local minimum. Conversely, if *ε* is too large, the estimates of the infectious population (from the `Simulate` module) can be invalid, leading to unstable updates. In-between, the parameter *ε* can guide the algorithm’s convergence. Ultimately, we solve Problem (𝒫) by replacing Equations (16)– (18) by Equations (32)–(34). We refer to this module as `Optimize` (**Î**, *ε*).

We iterate between the `Simulate` and `Optimize` modules, until convergence. Specifically, the algorithm terminates when both the value of the objective function and the estimates of the infected population remain unchanged from one iteration to the next. The pseudocode summarizing this approach is presented in Algorithm 1. Finally, note that the proposed algorithm depends on the initial feasible solution. We present next two approaches for generating this initial solution.

### 4.2 Feasible solution generation

We propose two interpretable heuristics for generating a feasible solution to (𝒫).

#### Randomized allocation

We generate a permutation of ℳ and use it as a randomized “ranking” of regions. In each time period *t*, we first assign to each region its minimum required allocation of vaccines, as determined by Equation (29)). Then, we allocate the remaining vaccines from the highest to lowest ranked region, until either no susceptible individual is left, or the region’s maximum capacity in time period *t* is reached. Within each region, we then allocate the available vaccines toeligible, susceptible individuals from highest to lowest risk class, according to the mortality rates.

##### Algorithm 1: Coordinate descent algorithm for DELPHI{V{OPT (Problem (𝒫)).

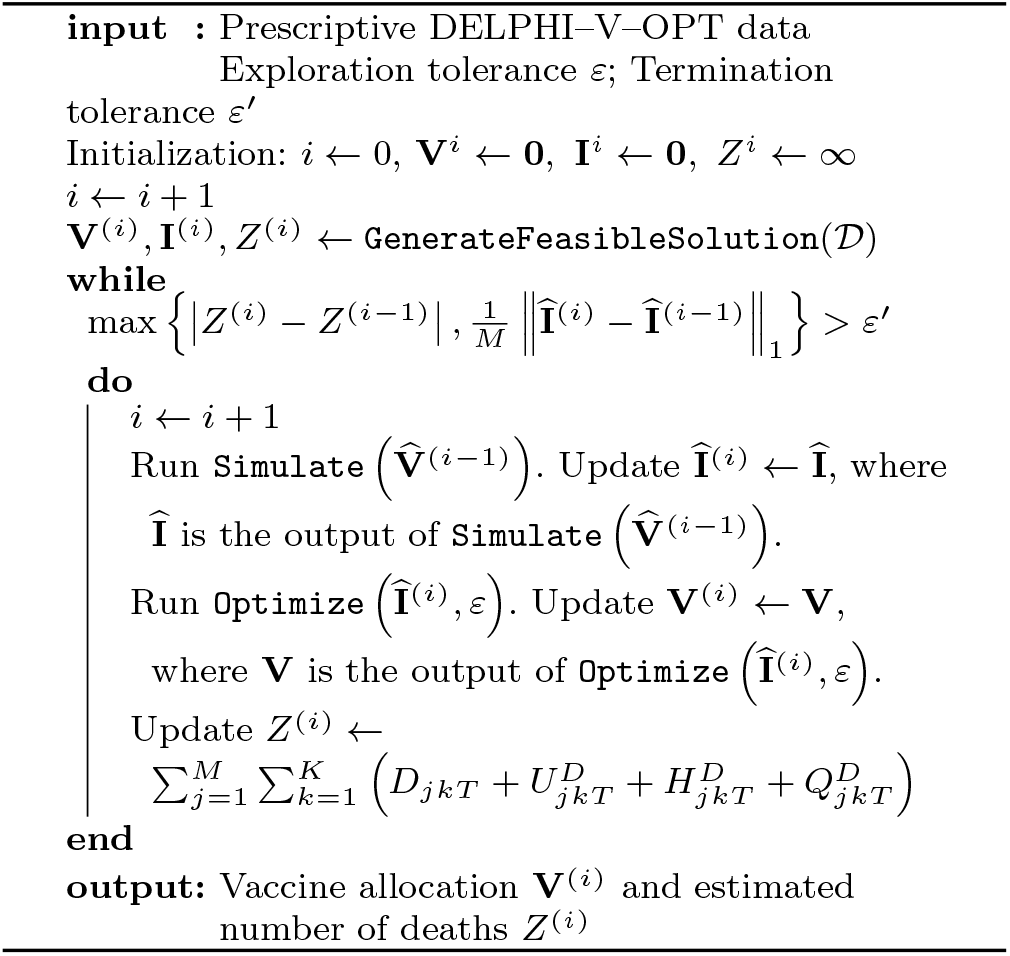

#### Prioritized allocation

This method provides an informed vaccine allocation strategy based on the output of the DELPHI–V model. In each time period, we assign vaccines to each region proportionally to the size of its eligible, susceptible population. Specifically, let 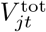 be the total number of vaccines allocated to region *j* in period *t*. We set:

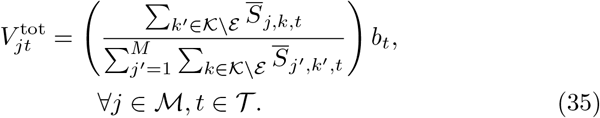

As with the randomized heuristic, we allocate the assigned vaccines within each region to eligible, susceptible individuals from highest to lowest risk class. By design, all constraints of Problem (𝒫) are satisfied. We refer to this initialization as the “warm start” heuristic.

### 4.3 Benchmark

Finally, we propose a realistic benchmark based on proportional allocation, defined as follows:

#### Proportional allocation

This method distributes vaccines proportionally to the (fixed) population size *N*_*jk*_ of each risk class *k* ∈ 𝒦\*ε* and region *j* ∈ ℳ. To ensure feasibility, we define the number of vaccines allocated to each subpopulation in each region as the minimum between the outcome of proportional allocation and the number of eligible people—although eligibility is rarely the limiting factor in practice. Mathematically, we have:

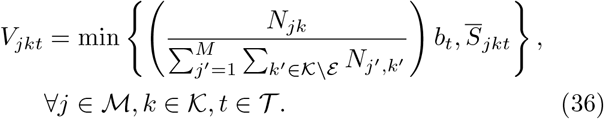

Again, by design, all constraints of Problem (𝒫) are automatically satisfied by this approach.

The proportional allocation is a sensible benchmark allocation of the vaccines, which ensures fairness across regions. Moreover, this approach is easily implementable, as it only relies on aggregate demographic information (e.g., census data) but does not involve the outputs of predictive modeling tools (such as DELPHI–V) on the dynamics of the pandemic or the effect of vaccinations. Accordingly, comparisons between our optimized solution and this benchmark provide estimates of the benefits of the proposed modeling and algorithmic approach for vaccine allocation.

## 5 Experimental setup

We implement the proposed model and algorithm in the United States. We consider allocation decisions at a state level, to mirror decisions faced by the Federal Government. Similarly, we limit our risk classes to six relatively coarse age groups: 0-9 years, 10-49 years, 50-59 years, 60-69 years, 70-79 years, and 80 years and above. These simplified risk classes facilitate the practical implementation of the allocation solution while capturing broad trends in mortality rates. We assume that no-one of age 9 and below or 80 and above can receive the vaccine for safety reasons, though this assumption can be re-evaluated once a vaccine is available.

We focus on two phases of the pandemic: (i) April 20, 2020 to July 15, 2020, and (ii) July 15, 2020 to October 15, 2020. This choice enables us to evaluate the benefit of the optimized vaccine allocation in retrospect (given ex post information on past dynamics of the pandemic), and to derive insights on optimal vaccine allocations given the future (at the time of the study) dynamics of the pandemic—obviously, the most salient outputs in practice. Moreover, the three-month periods provide a realistic planning horizon for largescale distribution of the vaccine.

### 5.1 Data sources

We calibrate the model using multiple data sources (Figure 3). First, we estimate the parameters of the DELPHI model (without vaccinations) independently for each state, using historical data on cases and deaths from the New York Times (2020). We obtain a granular population breakdown by age for each state from the US Census Bureau (2020b). We then run DELPHI (still, without vaccinations) to derive the initial number of susceptible, exposed and infected people (on April 20, 2020 and on July 15, 2020), which we distribute among the risk classes proportionally to their size. Figure 4 reports the predicted number of active cases over the April 20–July 15 period (prediction made on April 20) and over the July 15–October 15 period (prediction made on July 15), aggregated by geographic region (Northeast, Midwest, South and West).

**Fig. 3:**
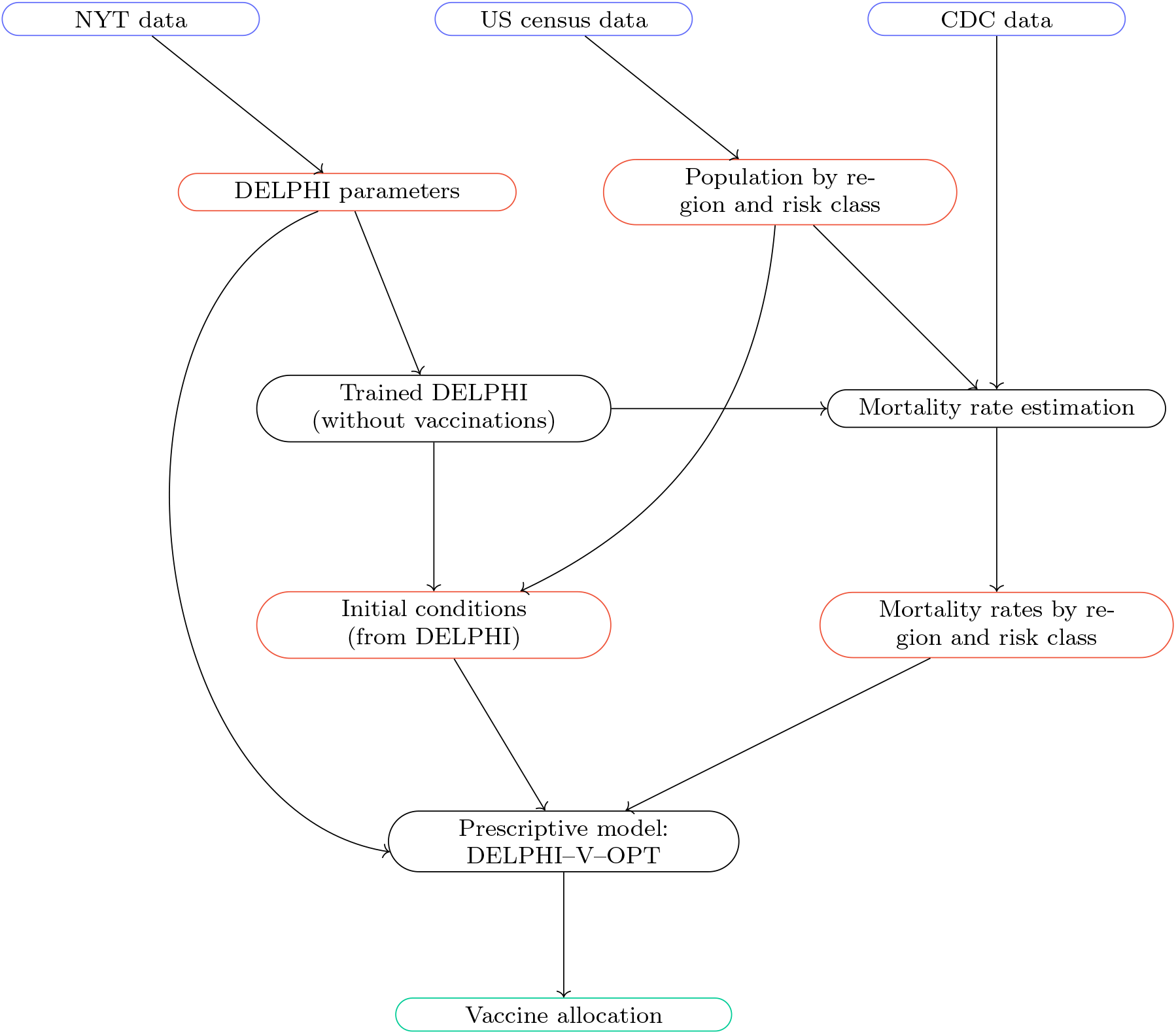
Simulation environment: raw data (blue), processed data (red), models (black) and outputs (green).

**Fig. 4:**
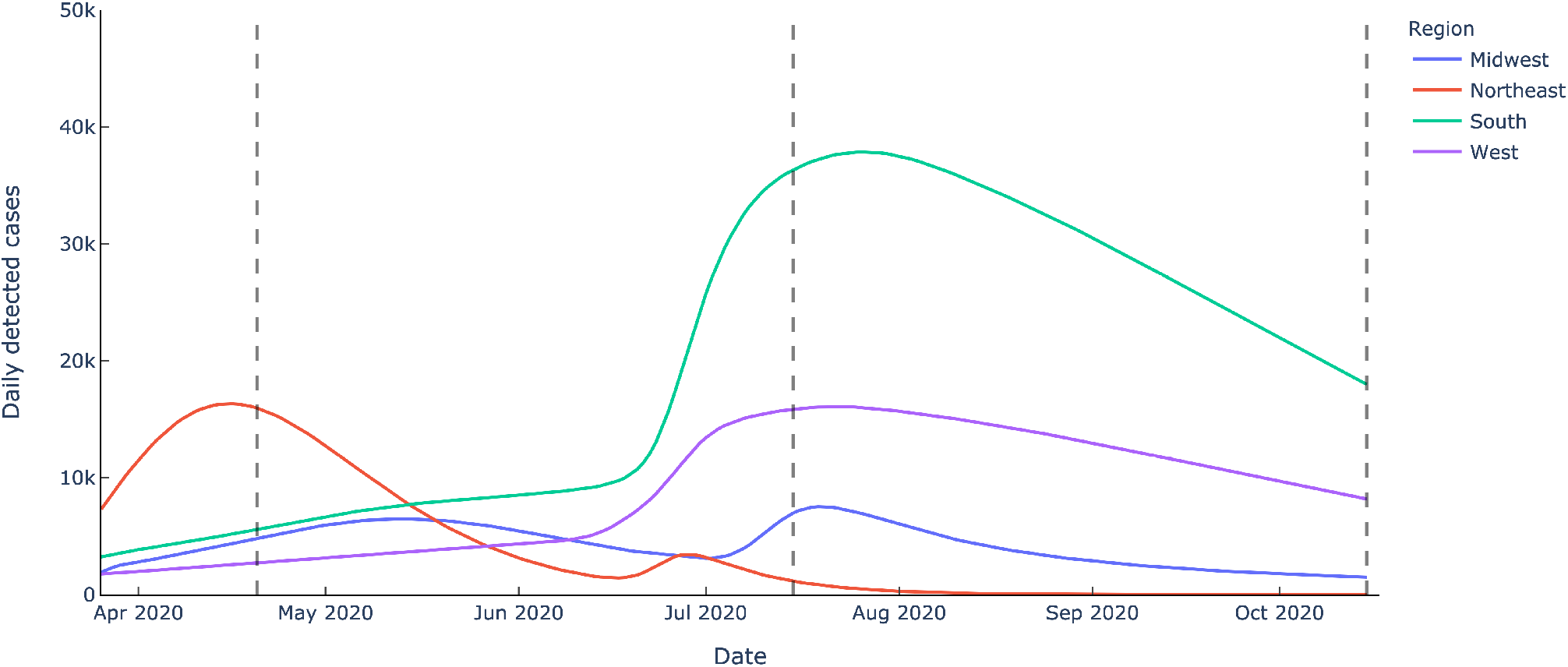
Daily detected cases predicted by DELPHI over the course of the pandemic, segmented by US geographic region and by simulation period.

The next input is an estimate of the mortality rate per region, risk class and time period. We make use of the data from the US Center for Disease Control (2020b), which report the total number of confirmed COVID-19 cases, hospitalizations and deaths by age group in the United States until the end of May 2020. In contrast, the DELPHI model fits a time-dependent mortality rate within each region. To the best of our knowledge, there exists no other data source for nationwide cases and deaths by age group. This leaves a discrepancy between the CDC estimates at the risk class level, and the DELPHI estimates at the level of each region and each time period. To reconcile these data, we employ an optimization procedure that interpolates the mortality rate per region, risk class and time period. We present this approach in the next section.

### 5.2 Mortality rate estimation

Our procedure to estimate mortality rates starts from two sets of inputs:

– **DELPHI predictions:** Let *Ĉ*_*j,t*_ denote the estimated number of new detected cases in region *j* and time period *t*. Let 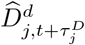 denote the number of deaths, where 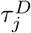 is the median death time after detection in region *j*. These quantities are aggregated across risk classes.
– **CDC data:** Let 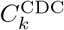 and 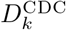 denote the total cases and deaths for risk class *k*. These quantities are aggregated across regions and time periods.

We define the reference mortality rate 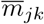 of risk class *k* based in region *j* as follows:

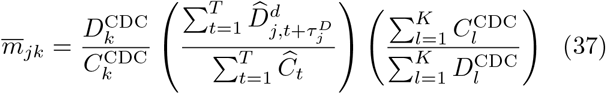

By design, this expression preserves the ratio of mortality rates between different risk classes from the CDC data, while correcting the mean reference mortality rate in each region across the planning horizon to be in line with the DELPHI projections.

We then estimate the mortality rate for each region *j* ∈ ℳ, risk class *k* ∈ 𝒦, and time period *t* ∈ 𝒯, denoted by *m*_*jkt*_. We also introduce additional decision variables *C*_*jkt*_ and 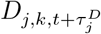, reflecting the number of detected cases and the number of future deaths in region *j* assigned to risk class *k* and time period *t*. Given that the fitting procedure is done separately in each region *j* ∈ ℳ, we decouple the problem at the region level—thereby considerably reducing the size of each problem instance. Specifically, we formulate the optimization problem given in Equations (38)–(45).

The first term in Equation (38) minimizes the squared relative error between the mortality rate estimates and their reference values (Equation (37)). The second term is a regularization penalty that minimizes deviations between the proportion of detected cases and the proportion *p*_*jk*_ of the population, in each risk class. The parameter *λ* trades off these two objectives (we use *λ* = 0.1 in our experiments). Equations (39)–(40) enforce that the total numbers of detected cases and deaths are consistent with the DELPHI predictions. Equation (41) defines the mortality rate as the ratio between the number of deaths and cases. Equations (42)–(43) ensure that the mortality rates are decreasing over time and monotonic as a function of the risk classes (as determined by the reference values). Finally, Equations (44)– (45) define the domain of the variables.

Note that the problem is non-linear due to the bilinear term in Equation (41). Yet, thanks to the decoupling at the region level, we can solve the problem using the quadratic solver in Gurobi 9.0 (Gurobi Optimization 2020), which addresses non-convexities using branching and cutting planes algorithms. In practice, a solution within a 1% optimality gap is generally obtained within minutes.

The output of this algorithm is an estimate of the mortality rate at the level of each state and each risk

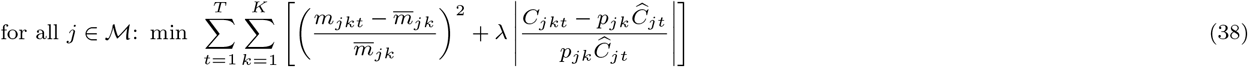

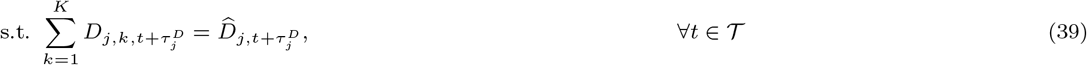

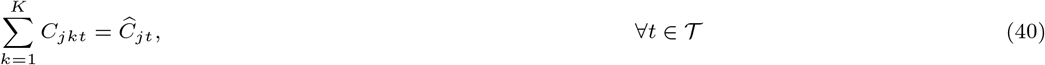

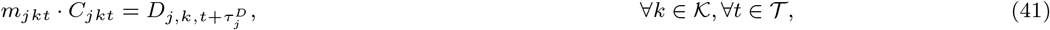

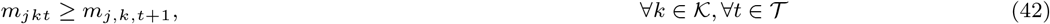

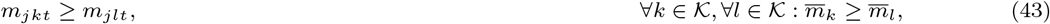

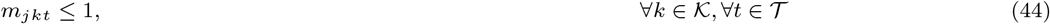

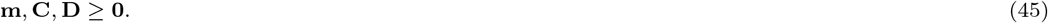

class, in each time period. We report aggregated statistics in Table 2.

**Table 2:** Calibrated monthly mortality rates, averaged by risk class.

| Month | 0-9 | 10-49 | 50-59 | 60-69 | 70-79 | 80+ |
| --- | --- | --- | --- | --- | --- | --- |
| April | 0.037% | 0.723% | 3.553% | 9.934% | 25.295% | 43.452% |
| May | 0.035% | 0.718% | 3.518% | 9.721% | 24.682% | 42.604% |
| June | 0.035% | 0.717% | 3.518% | 9.705% | 24.636% | 42.569% |
| July | 0.034% | 0.716% | 3.513% | 9.671% | 24.529% | 42.416% |
| August | 0.032% | 0.714% | 3.502% | 9.601% | 24.329% | 42.146% |
| September | 0.032% | 0.713% | 3.500% | 9.588% | 24.301% | 42.088% |
| October | 0.031% | 0.712% | 3.476% | 9.488% | 24.003% | 41.671% |

### 5.3 Implementation details

Our prescriptive DELPHI–V–OPT model relies on a forward difference scheme to replicate the evolution of the pandemic outbreak, for a given vaccine allocation. If such discretization is too coarse, the algorithm will introduce truncation errors in the estimates of the effects of vaccine allocations. If, however, such a discretization is too granular, the computational run times will be prohibitively long. We experimented with multiple values of *Δt* ∈ [0.01, 0.1, 0.5, 1, 5, 10] (units of days), and found *Δt* = 1 day to yield the best compromise between computational run times and modeling accuracy. It is also practical choice as it yields a day-by-day plan.

The coordinate descent scheme terminates when the change in both objective value and infection error lies below a pre-determined threshold. We set the termination tolerance to 500 (people), which provides a reasonable precision in light of the current number of deaths and active infections in the United States. Similarly, we set the exploration tolerance of Equation (34) to 500.

Regarding the DELPHI-V–OPT parameters, we set the maximum regional allocation fraction to 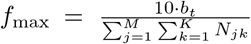, allowing each state to vaccinate a percentage of people up to one order of magnitude larger than the nation as a whole. We set *δ* = 0.1, meaning the amount allocated to any region can change by at most 10% of its maximum capacity from day to day. Next, we perform sensitivity analyses to assess the impact of vaccine effectiveness *β*, daily vaccine budget *b*_*t*_ and the minimum regional allocation fraction *f*_min_. The vaccine effectiveness and daily budget are key drivers of the overall impact of vaccinations, whereas the minimum regional allocation fraction determines how “fair” the allocation must be across regions. For simplicity, we consider only a constant daily vaccine budget and thus omit the subscript from hereon. Specifically, we consider values of vaccine effectiveness *β* equal to 0.3, 0.4, 0.5, 0.6, 0.7, and 1; a daily vaccine budget *b* of 100,000, 200,000, 300,000, 500,000, 1M, and 2M; and minimum regional allocation fractions *f*_min_ of 0, 0.1, 0.2, 0.3, 0.4, and 0.5. Obviously, a vaccine effectiveness of 100% is overly optimistic; yet it provides an ideal scenario against which to compare more realistic vaccine distribution strategies. The maximum budget is chosen to reflect the most aggressive objectives of vaccinating the entire US population by January 2021 (US Department of Health &Human Services 2020). Collectively, we thus obtain 6^3^ = 216 different sets of parameters.

All optimization models are implemented in Gurobi 9.0, with 2.3GHz processor and 4 cores. We use a barrier method to solve each linear program, with a barrier convergence tolerance of 10^−6^.

## 6 Experimental results

We now turn to the results of the modeling and algorithmic framework developed in this paper. We first show the convergence of the coordinate descent algorithm presented in Section 4 and its robustness to the initial conditions. We then show the benefits of vaccine allocation optimization, as compared to the proportional allocation benchmark, and we establish the robustness of these benefits to several model parameters. We conclude by describing the optimal vaccine allocation strategy to enhance interpretability of the optimization outputs.

### 6.1 Algorithmic convergence

Recall that the coordinate descent algorithm uses an iterative procedure to solve the nonlinear program (𝒫) (Algorithm 1). To show the algorithm’s convergence, Figure 5 plots the value of the objective function (i.e., number of deaths) at each iteration. We consider 11 initial solutions: ten randomized initializations and our warm start heuristic (prioritized allocation).

**Fig. 5:**
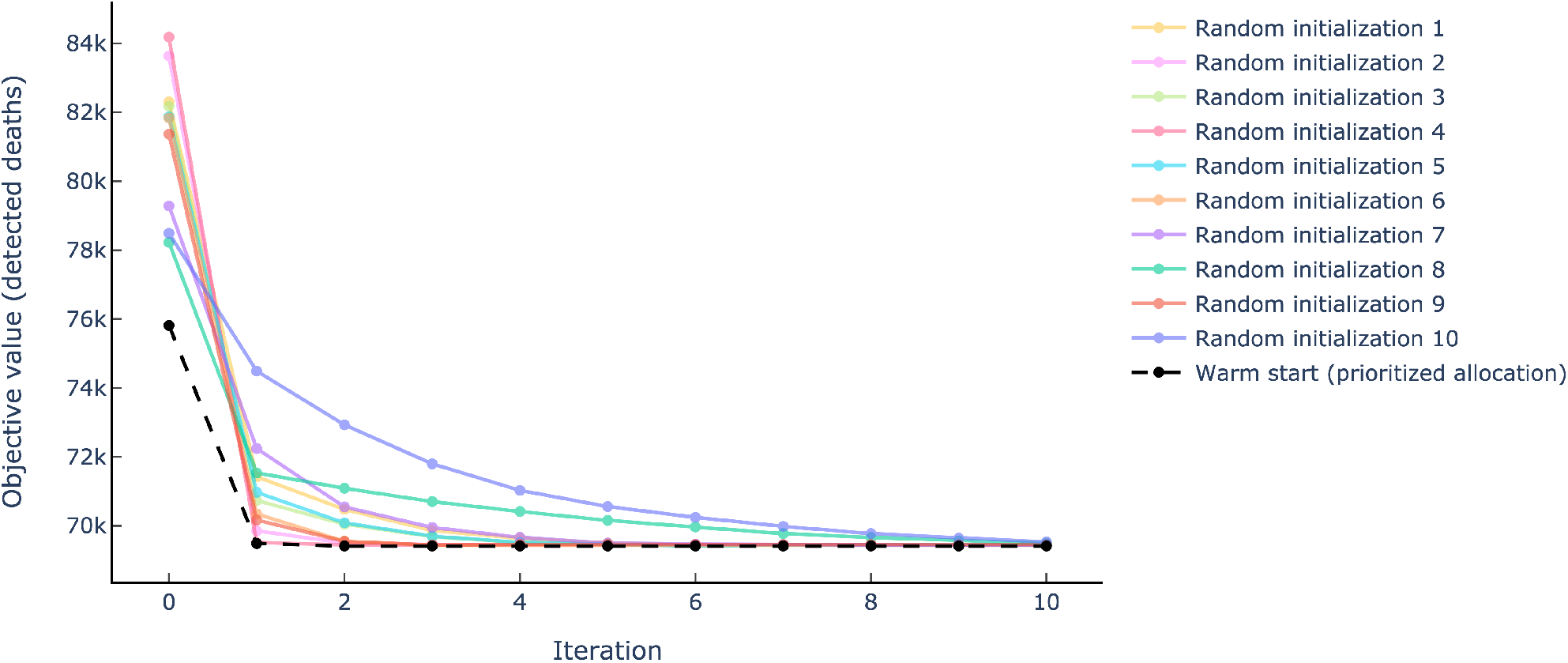
Convergence of Algorithm 1 using randomized and warm start initializations over the July 15–October 15 period (*β* = 0.6, *b* = 300k and *f*_min_ = 0.1).

Note that the algorithm converges in a small number of iterations, regardless of the initialization heuristic. In all 11 experiments, the algorithm reaches the same objective value, within the termination tolerance, in 10 iterations or less. These results show the benefits of our coordinate descent approach to solve the original non-convex optimization problem (𝒫), as well as its robustness to the choice of the initial solution. In contrast, a direct implementation of problem (𝒫) using the bilinear solver in Gurobi 9.0 failed to even find a feasible solution after 30 minutes of runtime.

Yet, the results also suggest that the warm-start prioritized allocation heuristic can help accelerate convergence. Indeed, when using the warm start solution, the algorithm converges in 2–4 iterations—a reduction of 60–80% as compared to the worst random initialization. In other words, the prioritized allocation heuristic can significantly reduce the computational requirements of the algorithm. These results underscore, in turn, the benefits of jointly using the outputs of the predictive DELPHI–V model in designing a good starting solution and iteratively solving the prescriptive DELPHI– V–OPT model to optimize vaccine allocation.

In addition to establishing the robustness of the optimal objective value, we also assess the stability of the optimized vaccine allocation solution. We define stability by using the mean absolute percent deviation (MAPD) of the cumulative immune population, averaged over time and across all 10 solutions obtained with randomized initializations (using the one obtained with the warm start as a reference point). Let 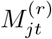 denote the number of immune people in region *j* in period *t* in the optimized solution obtained with randomized initialization *r* = 1, *…*, 10. Similarly, let 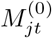 denote the number of immune people in region *j* in period *t* in the optimized solution obtained from the warm start initialization (recall that a strictly positive number of people receive a vaccine in each state in each time period due to the fairness constraints). The MAPD is then given as follows, for all *j* ∈ *J* :

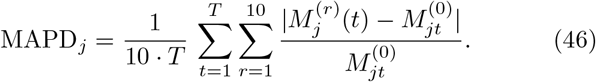

Table 3 reports aggregate statistics on the MAPD per US region. The results show that the number of allocated vaccines varies by only 1–5% on average across solutions. So, even though the initial allocations lead to different solutions, these solutions only differ mildly from each other. Coupled with the fact that these solutions achieve essentially the same objective function value (Figure 5), this indicates that the problem admits multiple close-to-optimal solutions and that the algorithm is able to find these different optima with different initialization points.

**Table 3:** Aggregate MAPD statistics by US region, over the July 15–October 15 period (*β* = 0.6, *b* = 300, 000 and *f*_min_ = 0.1).

| Region | Midwest | Northeast | South | West |
| --- | --- | --- | --- | --- |
| 10th percentile | 0.56 | 1.59 | 0.22 | 0.51 |
| Median | 1.19 | 5.26 | 1.19 | 1.25 |
| Mean | 3.16 | 5.22 | 2.25 | 2.26 |
| 90th percentile | 9.35 | 8.53 | 6.19 | 4.96 |

Figure 6 illustrates these patterns by plotting the number of allocated vaccines in each of four states over time. By design, these states are representative of all 51 states, with California having the most unstable solution (i.e., the largest MAPD), Oklahoma one of the most stable ones (i.e., the bottom quartile of MAPD), and Missouri and North Carolina falling in-between.

**Fig. 6:**
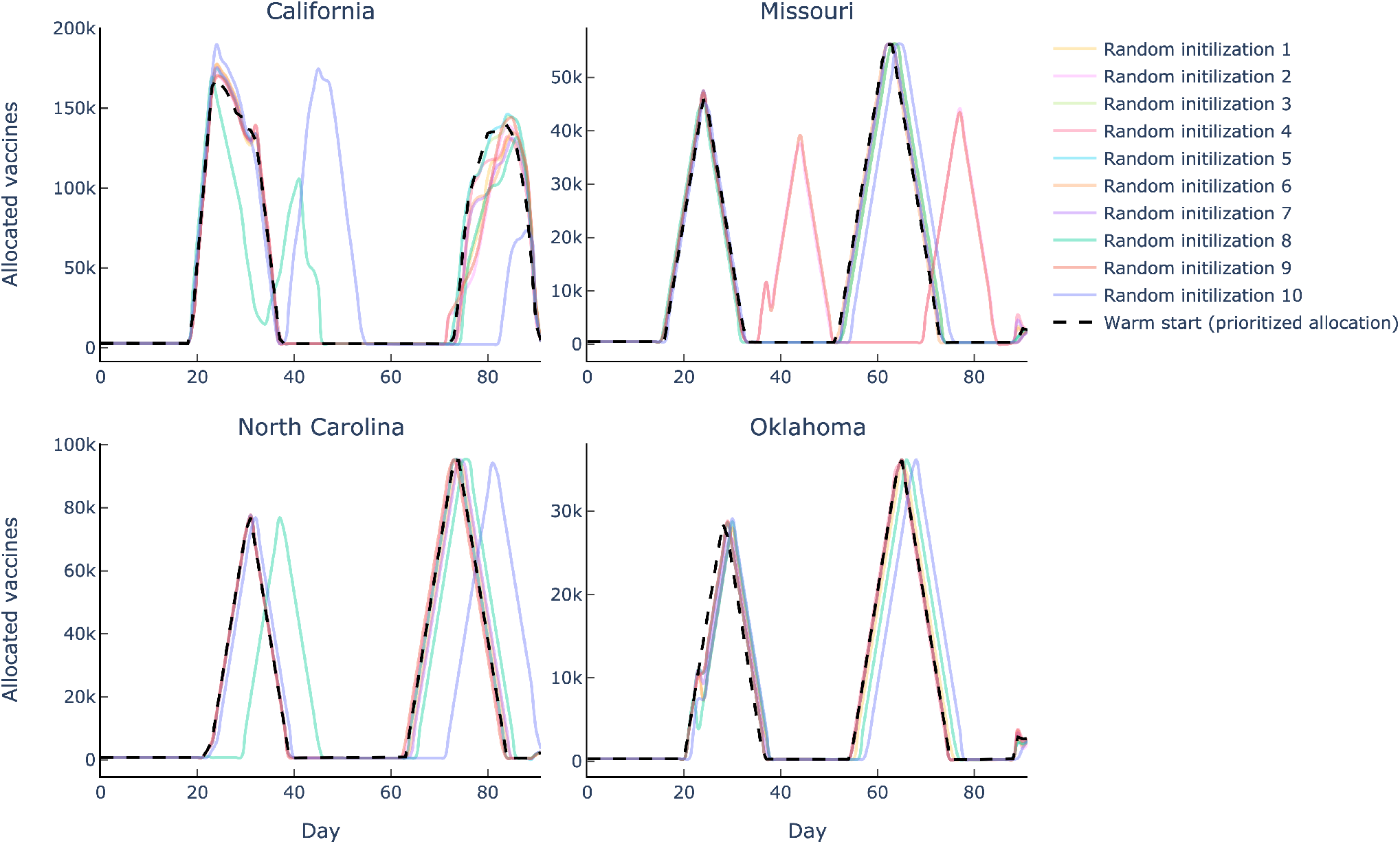
Solution stability, for four representative states, over the July 15–October 15 period (*β* = 0.6, *b* = 300, 000 and *f*_min_ = 0.1).

The main observation is that the solution is fairly stable across all solutions. Starting from Oklahoma, all 11 solutions essentially result in virtually the same vaccine allocation outcome, with a first peak after one month (for the risk class facing the highest risk) and another peak after two months (for second highest-risk class). For other states, all solutions are not completely identical, but remain highly similar. For instance, in Missouri, 10 out of the 11 solutions are virtually identical, whereas the last one simply delays the allocation by two weeks. Obviously, this delay in Missouri comes with earlier allocations elsewhere, resulting in the same overall impact on the death toll of the pandemic. In North Carolina, 9 solutions result in the same outcome, whereas one delays the first allocation peak and another one delays the second allocation peak. Finally, in California (again, the state with the largest average MAPD), solutions exhibit more variations but, still, the overall magnitude and dynamics of vaccine allocation remain similar across all 11 solutions.

From a practical standpoint, these results suggest that the proposed model can provide an effective decisionmaking tool to support vaccine allocation. Indeed, as pharmaceutical companies need to plan their operations well in advance of actual vaccine distribution, it is critical to get visibility into the future spatial-temporal dynamics of vaccine allocation. If we had obtained various “acceptable” solutions with the same overall number of deaths but widely different spatial-temporal patterns, it would have been challenging to use these data-driven results toward effective decision-making. However, this is not the case. Instead, the proposed model and algorithm yield highly stable recommendations that can be leveraged for vaccine allocation planning.

### 6.2 Benefits of vaccine allocation optimization

We now quantify the benefits of the proposed model and algorithm for vaccine allocation. To this end, we compare the outcomes of the optimized solution and the proportional allocation benchmark (which allocates vaccines proportionally to the size of each subpopulation). We evaluate both strategies using our predictive DELPHI–V model, which has been extensively validated as an effective tool to predict the future dynamics of the COVID-19 pandemic (Li et al. 2020).

Figure 7 shows the number of detected active cases and the cumulative number of detected deaths throughout the 90-day horizon. The main observation is that optimized vaccine allocation can result in significantly lower numbers of active cases and deaths than the benchmark allocation. The total number of deaths is reduced by 20–25% over the planning horizon (from approximately 70,000 to 50,000 deaths, in this case). This obviously represents a sizeable improvement, highlighting the benefits of the proposed analytics-based approach developed in this paper.

**Fig. 7:**
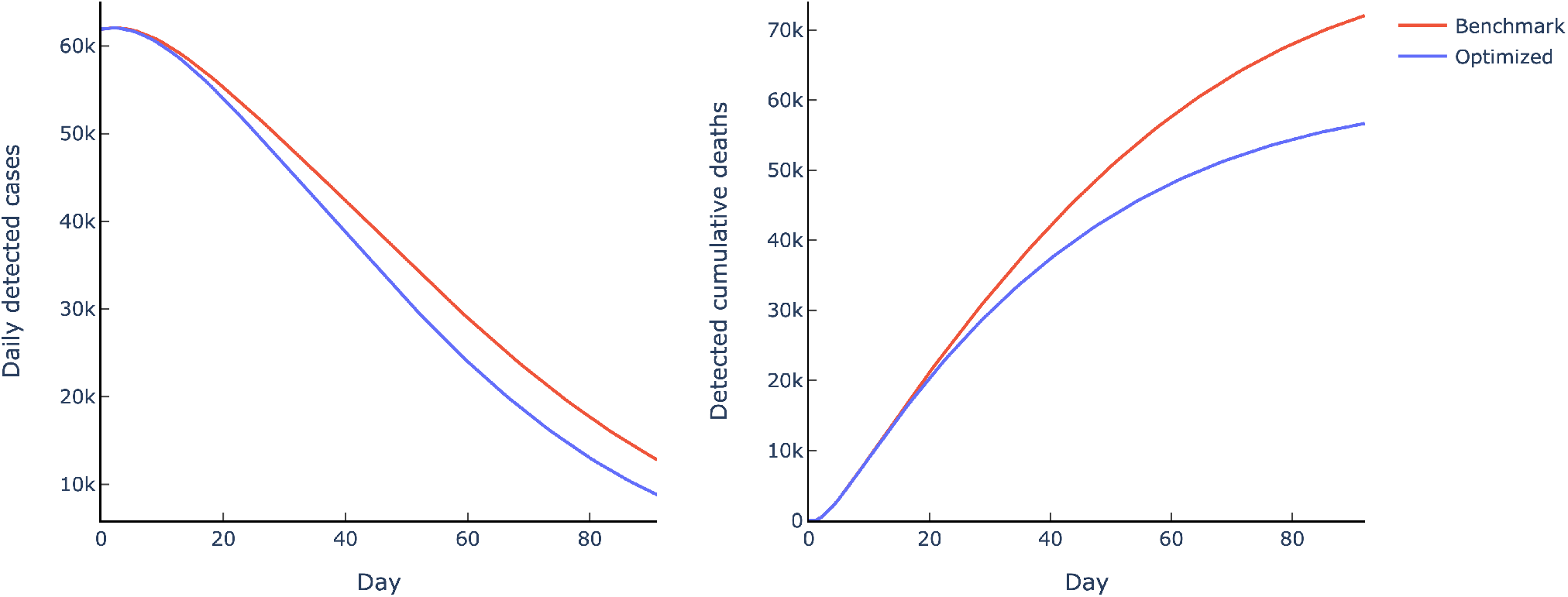
Projected number of daily detected cases and cumulative deaths for optimized vs. benchmark vaccine allocations, over the July 15–October 15 period (*β* = 0.6, *b* = 1, 000, 000 and *f*_min_ = 0.1)

Two additional observations from this figure are noteworthy. First, the number of active cases is also reduced thanks to the optimized vaccine allocation strategy. The number of active cases is projected to begin declining over the July 15–October 15 period under both allocation strategies (due to the governmental restrictions in place) but the decline is faster and stronger under the optimized vaccine allocation. This was not an obvious outcome, since the model’s objective is to minimize the total number of deaths—not the number of cases. Yet, the proposed model ends up prioritizing vaccines where infections are highest, precisely to shield the most vulnerable subpopulations where they are the most exposed to the disease. This allocation strategy results in a lower number of cases overall. This observation suggests that the model does not have disproportionate adverse consequences on low-risk subpopulations—either states with fewer cases or low-risk classes.

Second, the benefits of the optimized vaccine allocation strategy come with a time lag. In this instance, the distribution of vaccines starts on Day 1 (July 15), but the optimized allocation results in a markedly lower number of deaths starting from Day 30 (mid August onward). This is due to the non-linear SEIR dynamics that govern the DELPHI–V model: vaccines initially reduce the susceptible population, which results in lower numbers of newly infected people, of active cases, and, ultimately, of deaths. Again, this result has important practical implications, underscoring the role of data and long-term dynamics in vaccine allocation.

Figure 8 plots the sensitivity of the reduction in the number of deaths resulting from the optimized vaccine allocation strategy against the proportional allocation benchmark, as a function of vaccine effectiveness and the daily budget of vaccines. Note, first, that the benefits of the proposed model and algorithm increase with vaccine effectiveness. This is expected, as a higher vaccine effectiveness increases the overall impact of all vaccine allocation strategies (in the extreme example where *β* = 0, all allocations have the same performance). Moreover, the benefits of optimized vaccine allocation exhibit a unimodal relationship with the daily budget. Again, all allocations have the same performance when no vaccine is available (*b* = 0). As the budget increases, the decisions of *who* should receive vaccines and *when* anyone should receive a vaccine become increasingly complex. As a result, the optimized allocation strategy has a positive and significant impact on the dynamics by curbing the spread of the disease and the resulting death toll more significantly than the benchmark allocation. However, after a certain point, the budget is so high that both allocations perform very well, thus reducing the edge of the optimized strategy as compared to the benchmark. Yet, in a reasonable range of daily budgets (from 200,000 to 2,000,000 vaccines available in the United States per day), the proposed allocation strategy can have a very significant impact on the pandemic, reducing the number of fatalities by 10–25%, or 10,000–20,000 deaths in the United States over a three-month period.

**Fig. 8:**
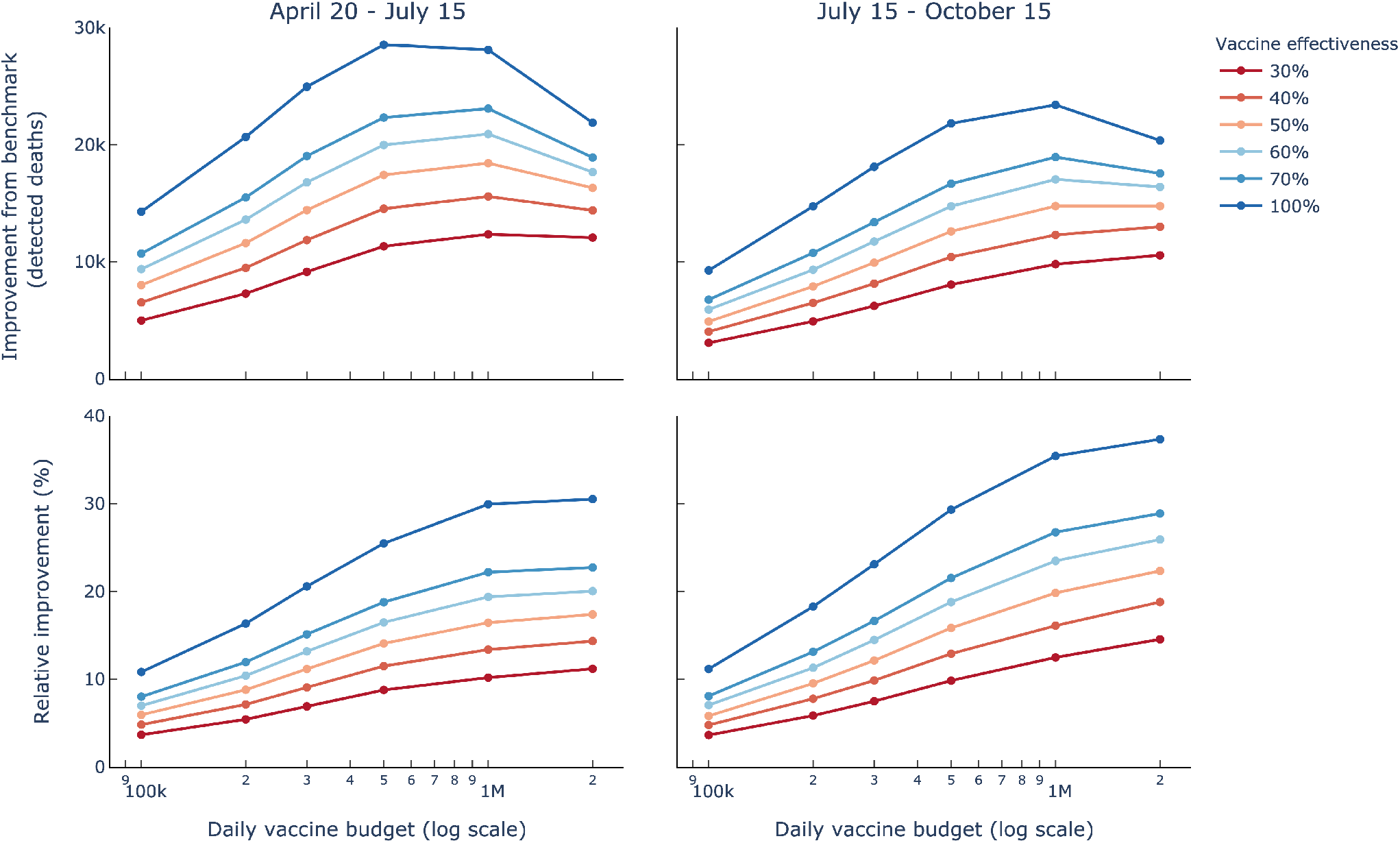
Absolute (top) and relative (bottom) improvement from the optimized vaccine allocation, over the July 15–October 15 period, as a function of vaccine effectiveness *β* and the daily budget *b* (*f*_min_ = 0.1).

We conclude by investigating the price of fairness in the optimized vaccine allocation strategy. Recall that our formulation ensures that a fraction of at least *f*_min_ of the eligible susceptible population receives a vaccine in each state. Figure 9 plots the relative improvement, as compared to the proportional allocation benchmark, as a function of *f*_min_, for different daily budgets. Note that the relative improvements decline as the fairness constraints become more stringent—since stronger fairness requirements reduce the remaining flexibility to allocate vaccines freely. Yet, this decline is very moderate. These results suggest that the edge of the optimized vaccine allocation is highly robust to the fairness constraints that need to be imposed in practice.

**Fig. 9:**
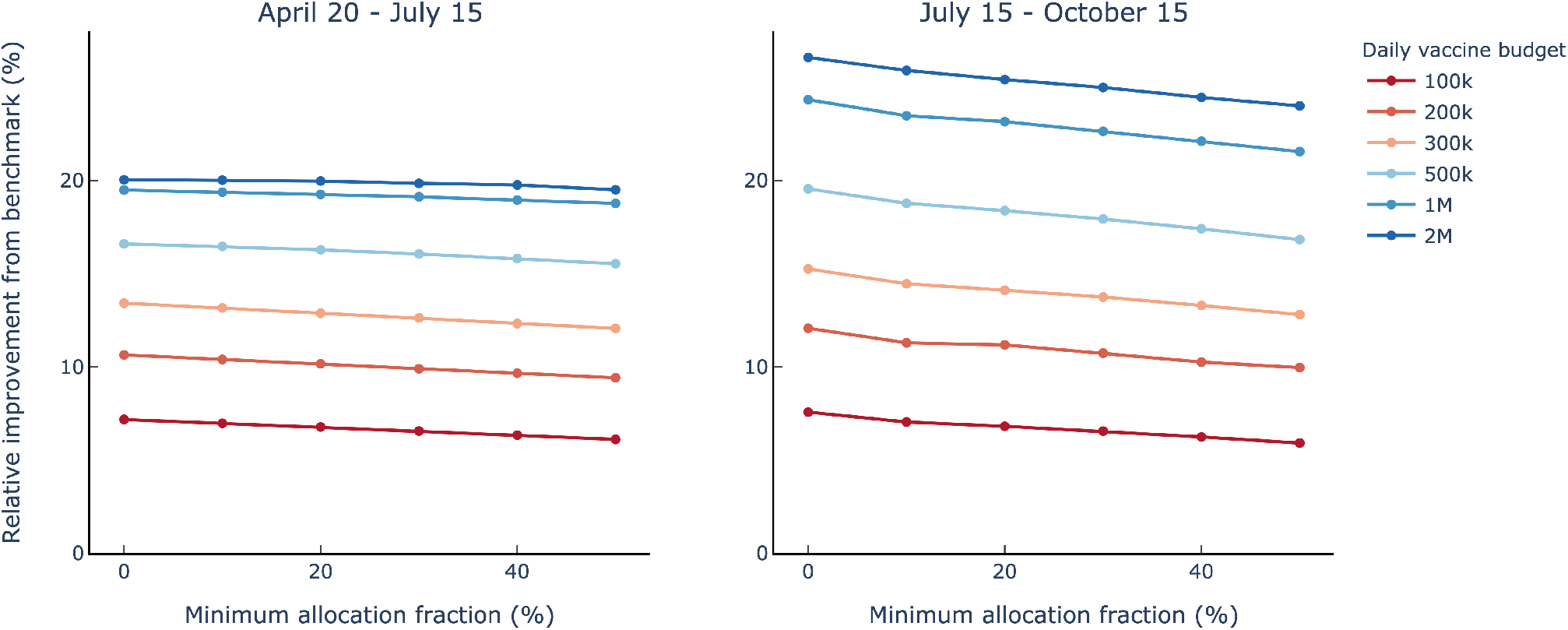
Relative improvement from the optimized vaccine allocation, over the July 15–October 15 period, as a function of the fairness parameter *f*_min_ and the daily budget (*β* = 0.6).

In summary, optimized vaccine allocation can have a significant effect on the death toll of the COVID-19 pandemic. Stated differently, the ultimate impact of the pandemic does not only depend on *when* a vaccine can be produced, on the *effectiveness* of the vaccine and on the *number* of vaccines available, but also on *how* these vaccines are allocated across the population. Our results demonstrate the potential of the proposed approach to optimize vaccine allocation accordingly.

### 6.3 Robustness of vaccine allocation

An important challenge when planning for vaccine distribution lies in the significant uncertainty regarding the state of the epidemic when a vaccine becomes available (i.e., the initial conditions in DELPHI–V), and the future dynamics of the epidemic (i.e., the various parameters and the very structure of DELPHI–V). We assess the robustness of the optimized vaccine allocation when model parameters are subject to change. Specifically, we first optimize vaccine allocation for the baseline parameter values, described in Section 5. We then perturb two key parameters in the model: the infection rate, which governs the transitions from susceptible to infected, and the mortality rate, which governs the fraction of the infected population that will die from the disease. For both parameters, we introduce a random perturbation for each region following a uniform distribution, with increasing percent-wise maximum deviations from the baseline value (the perturbation varies per region but not over time, so both rates remain constant through the planning horizon). Last, we simulate the dynamics of the pandemic using DELPHI–V with the new parameters, under the optimized allocation (obtained with the baseline parameters) and the benchmark allocation.

Figure 10 shows the distribution of the relative improvement resulting from the optimized vaccine allocation, as compared to the proportional allocation benchmark. In this instance, the optimized vaccine allocation strategy reduces the number of deaths by 14.5%, under the nominal parameter values. Obviously, these benefits vary with the infection and mortality rates. Yet, results show that the benefits of optimization remain highly robust to variations in the DELPHI–V parameters. Even for the largest deviations in the infection rates (by up to 50% from the baseline values estimated from historical data), the worst-case benefits still amount to nearly 10%, which, again, represents a highly significant reductions in the death toll of the pandemic. In fact, the variations can go either way: in over half the simulations, the relative benefits of the optimized allocation are even higher under the perturbed parameter values than under the nominal values.

**Fig. 10:**
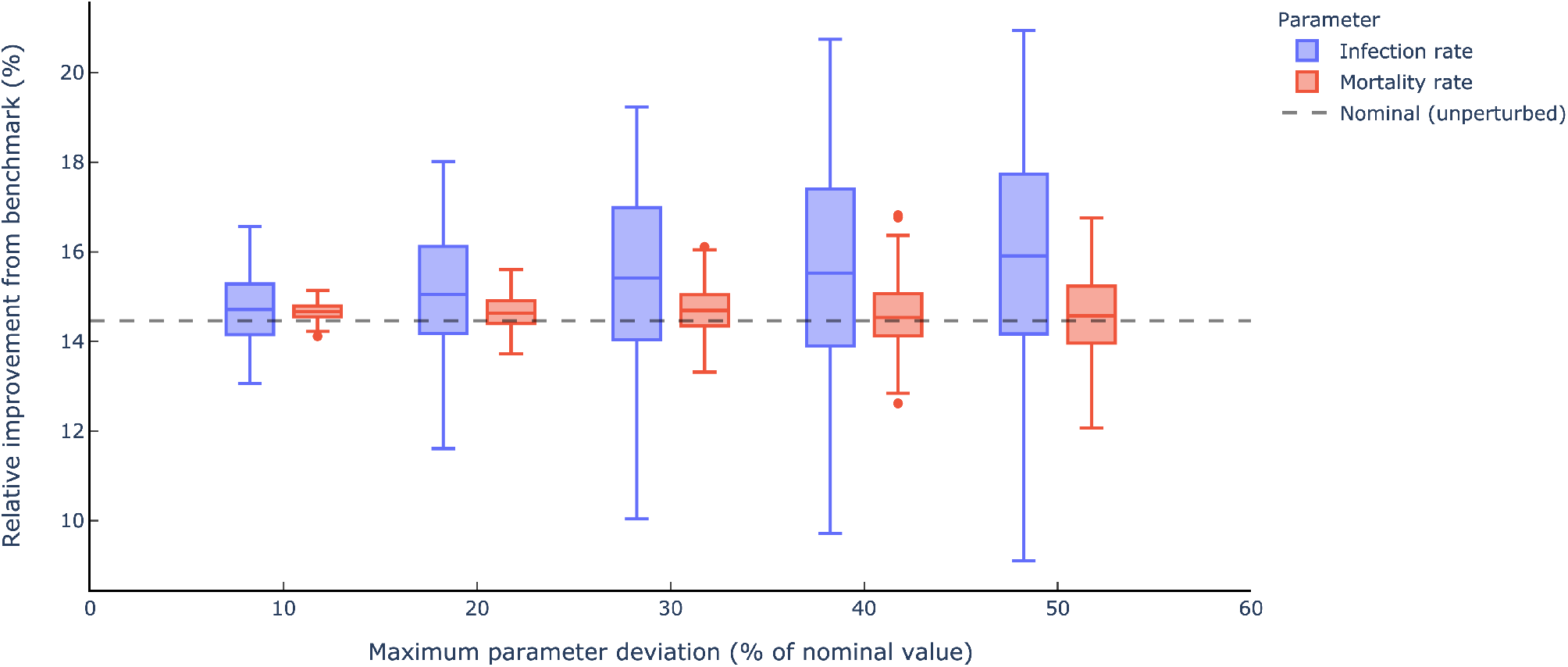
Relative improvement from optimized allocation, for increasing large perturbations in the infection and mortality rates, over the July 15–October 15 period (*β* = 0.6, *b* = 300, 000 and *f*_min_ = 0.1).

Note, finally, that the infection rate has a more significant impact on the relative benefits of optimized vaccine allocation than the mortality rate. This suggests that the impact of the pandemic depends mainly on how vaccines can curb infections at the upstream level—as opposed to mortality at the downstream level. This, again, illustrates the effects of the non-linear SEIR dynamics on the spread of the disease, and how we can leverage an epidemiological model such as DELPHI–V to allocate resources strategically in order to combat the pandemic most effectively.

### 6.4 Interpretability of optimized vaccine allocation

At this point, we have established the benefits of the proposed optimization approach to vaccine allocation, as opposed to a benchmark based on proportional allocation. But who should receive vaccines in priority at various stages of the pandemic? To address this question, Figure 11 illustrates the optimized vaccine allocations for two states: South Carolina and Oregon.

**Fig. 11:**
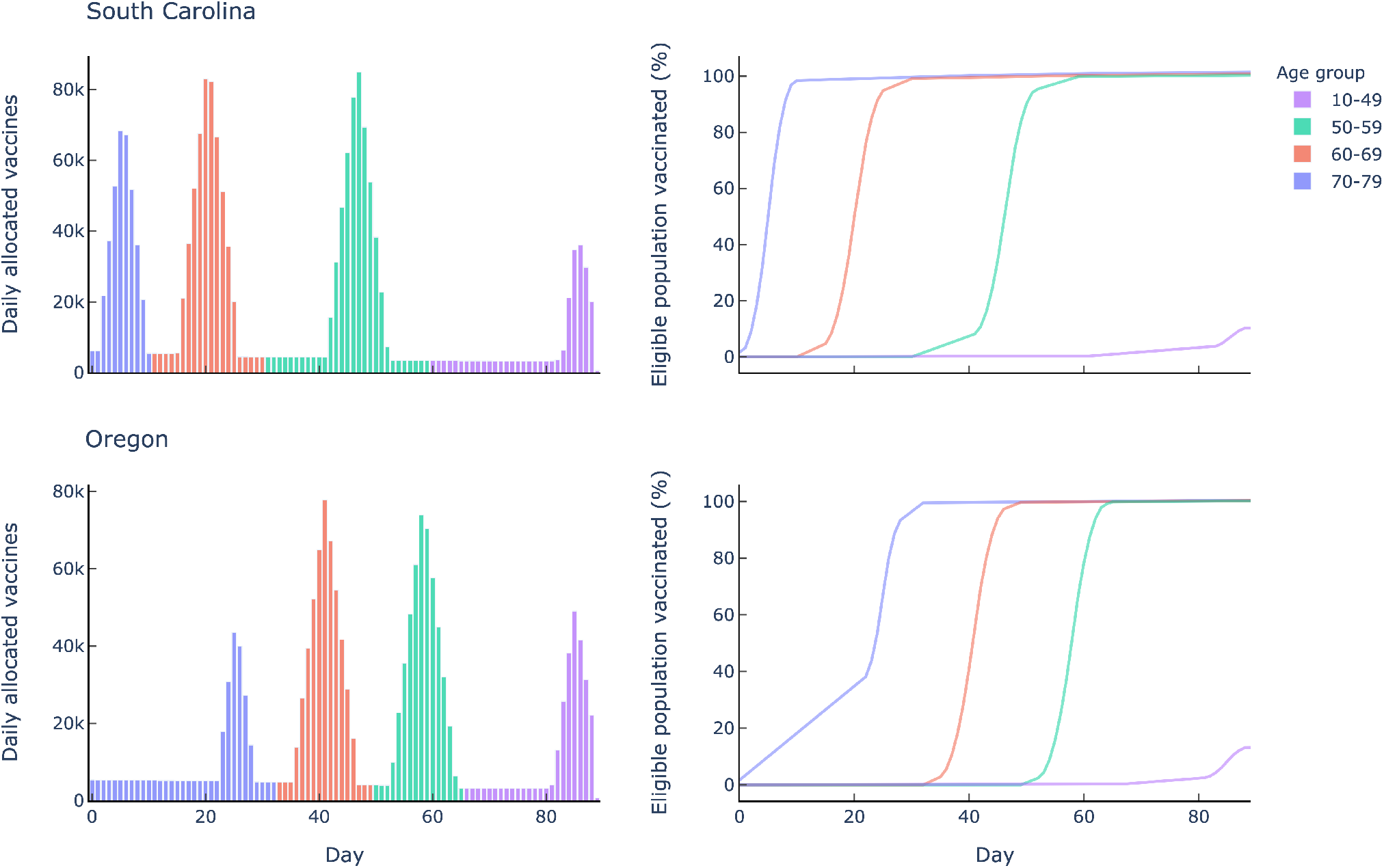
Optimized vaccine allocation in South Carolina and Oregon over the July 15–October 15 period (*β* = 0.6, *b* = 1, 000, 000 and *f*_min_ = 0.5).

The observations fall into two categories: the allocation across different states and the allocation across different age groups. First, vaccines are allocated to different states at different stages of the pandemic, based on local characteristics such as the number of active cases and the size of the susceptible population. As a result, the optimal vaccine allocation exhibit spatial-temporal variations. In the example pictured here, South Carolina gets prioritized ahead of Oregon (though, from the fairness constraints, all states receive a small but steady supply of vaccines throughout the planning horizon). Second, within each state, vaccines get allocated by order of risk priority, captured here by the age groups. In each state, vaccines get allocated to the older population first, until 100% of people aged 70–79 receive a vaccine (recall that we exclude the population aged 80 and over for safety reasons). Then, the second wave of vaccines get allocated to people aged 60–69, the third wave to people aged 50–59, and so on.

These results highlight the two main drivers of optimal vaccine allocation: (i) the near-term dynamics of the pandemic in every state and (ii) the risk level of each risk class within each state. All else equal, vaccines should be allocated in priority to the states that are most exposed to the pandemic and, within each state, vaccines should be allocated first to people facing the highest risk (this may also include healthcare and other essential workers, although these are not captured explicitly by our model). However, this leaves one question open: who to prioritize between a medium-risk population in a “hot spot” and a high-risk population in a state that remains relatively spared by the pandemic?

Figure 12 provides a visualization to answer this question. The vertical axis reports the age groups, from youngest (top) to older (bottom). The horizontal axis reports the 51 states, ordered by the number of predicted deaths per capita under the DELPHI model (without vaccinations)—an indicator of the near-term impact of the disease in each state. In other words, states are ordered from highest risk (left) to lowest risk (right). Importantly, the state “ranking” is determined by the predictive DELPHI model, as opposed to the prescriptive DELPHI–V–OPT model—thus ordering states without any consideration of the optimization outputs.

**Fig. 12:**
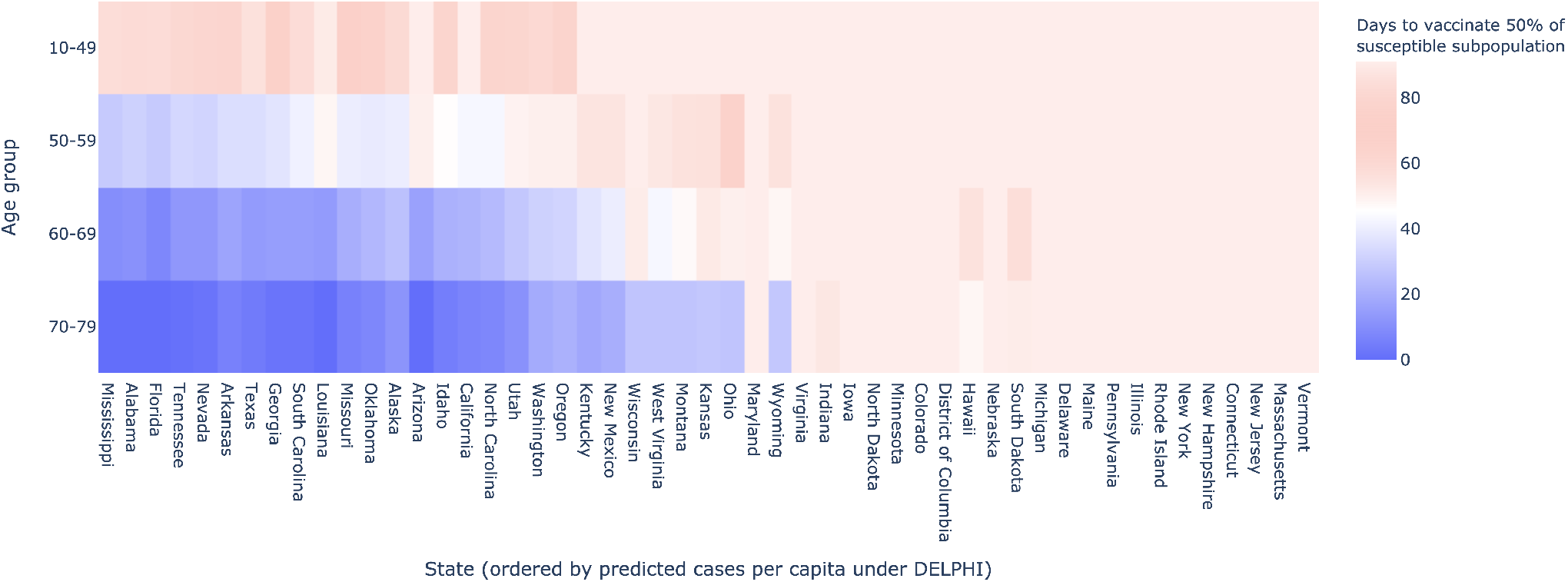
Time by which 50% of each subpopulation receives a vaccine per state and age group, over the July 15–October 15 period (*β* = 0.6, *b* = 1, 000, 000 and *f*_min_ = 0).

**Fig. 13:**
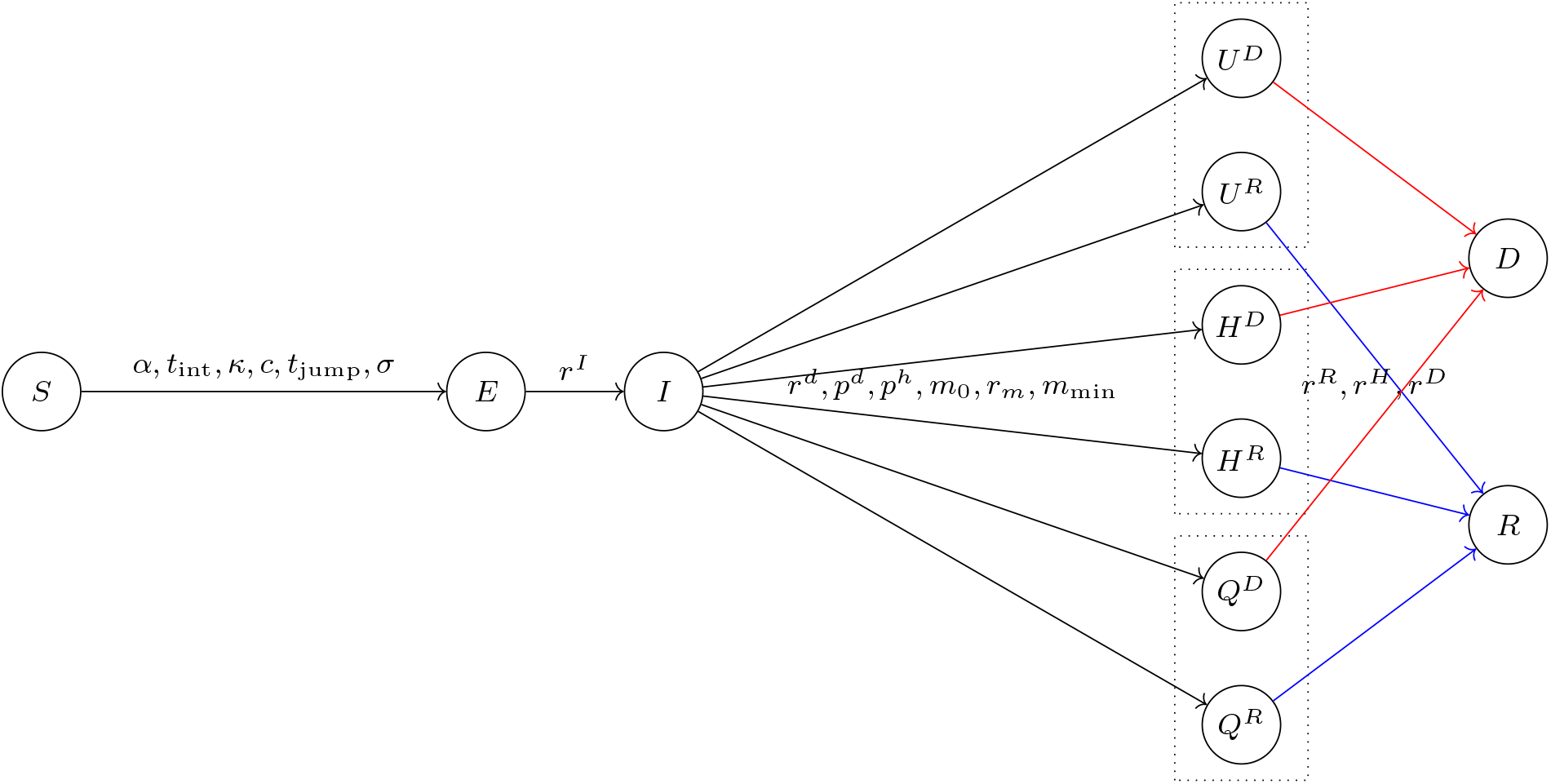
The DELPHI model.

This figure shows that the optimal solution to the trade-offs underlying vaccine allocation is not obvious, and governed by a number of interdependent dynamics. Clearly, the optimized solution prioritizes states that are more exposed to the pandemic (on the left-hand side) and the most vulnerable subpopulations (at the bottom). However, there is no clear ordering between these two objectives. Instead, the proposed algorithm optimizes vaccine allocation by balancing factors specific to each state and factors specific to each risk class.

## 7 Conclusion

This paper has presented a new prescriptive approach to vaccine allocation in response to the COVID-19 pandemic. The proposed approach starts with a state-of-the-art epidemiological model called DELPHI, which extends traditional SEIR models by capturing dynamics specific to COVID-19 (under-detection, governmental policies, societal response, and declining mortality rates). This paper has first proposed an extension, named DELPHI–V, which captures the effects of vaccinations and reflects the disaggregated impact of COVID-19 on mortality across risk classes (e.g., age groups). Then, this paper has embedded the predictive DELPHI–V model into an optimization model, termed DELPHI–V– OPT, to inform vaccine allocation. DELPHI–V–OPT is formulated as a bilinear optimization model, and solved using a tailored algorithm based on coordinate descent.

Experimental results using real-world data in the United States suggest that the proposed optimization approach can yield significant benefits, as compared to a benchmark allocation that distributes vaccines proportionally to each subpopulation size. Specifically, the DELPHI–V–OPT model can reduce the death toll of the COVID–19 pandemic by an estimated 10–25% (over the most likely parameter range), or 10,000–20,000 deaths in the United States alone, over a three-month planning horizon. To achieve these benefits, the DELPHI– V–OPT model trades off allocating vaccines in the “hot spots” of the pandemic vs. allocating vaccines to the most vulnerable risk classes. The outputs of the DELPHI– V–OPT model can provide critical decision-making support to pharmaceutical companies and governmental agencies as they are currently planning for widespread vaccine distribution—in fact, the model is currently being used by a major US vaccine manufacturer.

Obviously, the optimization approach developed in this paper is not without limitations. For instance, our experiments have only partitioned the population according to age groups, thus ignoring other objectives such as prioritizing allocations to healthcare workers, other essential workers, or patients with comorbidities. Moreover, our epidemiological model does not capture heterogeneity in population mixing across subpopulations (e.g., interactions may be more intense in urban areas and between young people than in rural areas and between older populations). Finally, our methodology relies on a time discretization approximation and a coordinate descent approach, which do not yield theoretical guarantees on solution quality.

Whereas these limitations undoubtedly motivate further research, this paper lays the first data-driven brick on the optimal allocation of COVID-19 vaccines. At a time where the race to a vaccine is going full speed, the results from this paper highlight the critical role of vaccine allocation to combat the pandemic. Obviously, it is essential to make every effort possible to develop a vaccine as early as possible, to design a vaccine that is as effective as possible, and to get ready to manufacture as many vaccines as possible. But this paper shows another lever that can be pulled to curb the effect of the pandemic: strategically managing vaccine stockpiles to prevent the spread of the pandemic at the upstream level and to mitigate its impact at the downstream level.

## Data Availability

All data in this model are from the public JHU repository for COVID-19.

## A DELPHI parameter estimation

Recall that DELPHI is a compartmental epidemiological model that comprises 11 states. Its dynamics are governed by a system of ordinary differential equations (ODE). These ODE follow are similar to Equations (3)–(13), with three exceptions: (i) there is no vaccination term *V*_*k*_(*t*); (ii) there is no immune state *M*_*k*_; and (iii) the population is not disaggregated by risk class, so there is no index *k*. For completeness, we provide the model structure in Figure 13 and the system of ODE in Equations (47)–(57).

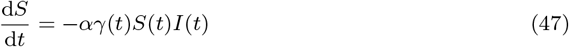

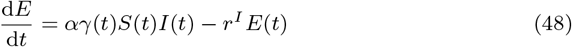

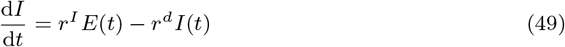

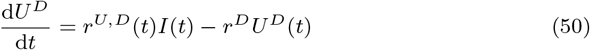

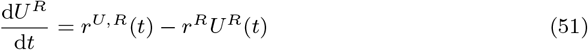

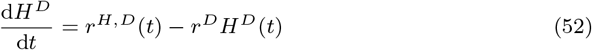

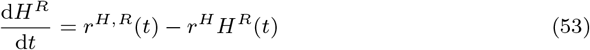

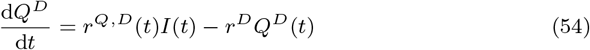

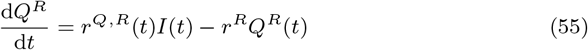

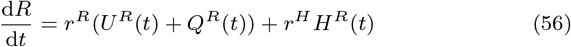

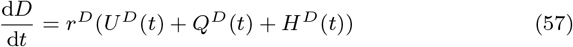

Recall that we parametrize the governmental and societal response function *γ*(*t*) as follows (Equation (1)):

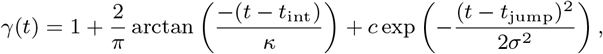

where *t*_int_ and *κ* denote the start time and the strength of the first-wave response, *t*_jump_ denotes the time of re-implementation of the restrictions, and *σ* controls the duration of the second wave.

Similarly, we parametrize the mortality rate *m*(*t*) as follows (Equation (2)):

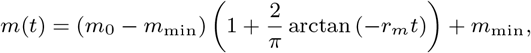

Next, we write the disaggregated detection, hospitalized and death rates as follows:

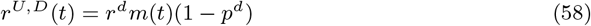

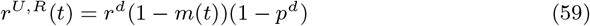

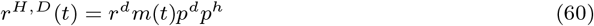

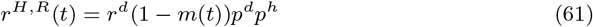

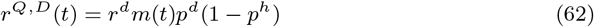

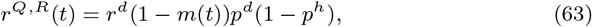

where *p*^*d*^ denotes the detection probability and *p*^*h*^ denotes the proportion of hospitalizations.

Ultimately, the system dynamics are governed by 16 explicit parameters, shown on the appropriate arrows in Figure 13. We introduce two additional parameters to account for the (unknown) intial population in the infected (*I*) and exposed (*E*) states. We therefore obtain 18 parameters per region.

To limit the amount of data needed to train this model, we fix 7 parameters using clinical data from a meta-analysis of over 190 papers on COVID-19 available at the time of the model’s creation (Bertsimas et al. 2020):

– *r*^*d*^ (detection rate) is equal to 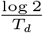, where *T*_*d*_ is the median time to detection (fixed to be 2 days) (Wang et al. 2020).
– *r*^*I*^ (infection rate leaving incubation phase) is equal to 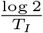, where *T*_*I*_ is the median time to leave incubation (fixed at 5 days) (Lauer et al. 2020).
– *r*^*R*^ (recovery rate of non-hospitalized patients) is equal to 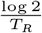, where *T*_*R*_ is the median time to recovery of non-hospitalized patients (fixed at 10 days) (Hu et al. 2020, Kluytmans et al. 2020).
– *r*^*H*^ (recovery rate of hospitalized patients) is equal to 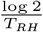, where *T*_*RH*_ is the median time to recovery under hospitalization (fixed at 15 days) (Liu et al. 2020, Grein et al. 2020).
– *m*_min_ (minimum mortality rate) is set to 1%, the lowest estimates of general COVID-19 mortality (Baud et al.2020). Note this quantity is independent from the rate of death.
– *p*^*d*^ (percentage of infectious cases detected) is set to 20% (Wang et al. 2020, Krantz and Rao 2020, Niehus et al.2020).
– *p*^*h*^ (percentage of detected cases hospitalized) is set to 15% (Arons et al. 2020, Xu et al. 2020).

We then fit the remaining 11 parameters from historical data:

– *α* (baseline infection rate)
– *t*_int_ (timing of governmental and societal response)
– *κ* (strength of governmental and societal response)
– *c* (strength of the second-wave resurgence)
– *t*_jump_ (time of the resurgence)
– *σ* (length of the resurgence)
– *m*_0_ (initial mortality rate)
– *r*_*m*_ (decay rate of mortality)
– *r*^*D*^ (death rate, i.e., speed at which a dying patient dies)
– *k*_1_ (initial infected population)
– *k*_2_ (initial exposed population)

Specifically, we fit these parameters by minimizing a weighted Mean Squared Error (MSE) metric. Let *DC*(*t*) and *DD*(*t*) denote the number of reported total detected cases and deaths on day *t*, defined as follows:

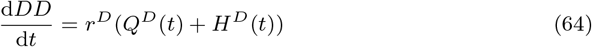

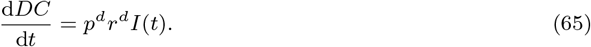

Then, the loss function for a training period of *T* days is defined as:

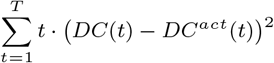

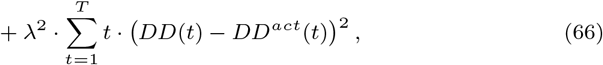

where *DC*^*act*^(*t*) and *DD*^*act*^(*t*) are actual numbers of detected cases and deaths. The factor *t* gives more prominence to more recent data, as recent errors are more likely to propagate into future errors. The weight 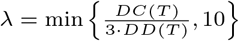 balances the fitting between detected cases and deaths. We only include historical data starting when the area recorded more than 100 cases; this allows us to exclude sporadic outbreaks that are not epidemics. We use non-convex optimization methods, including trust-region methods (Byrd et al. 2000) and the Nelder-Mead method (Lagarias et al. 1998), to minimize this loss function.

## Notes

### Competing Interest Statement

The authors have declared no competing interest.

### Funding Statement

None.

